# Backfill Bayesian Ordered Lattice Design for Phase I Clinical Trials

**DOI:** 10.64898/2026.04.02.26350086

**Authors:** Gi-Ming Wang, Curtis Tatsuoka

## Abstract

The Bayesian Ordered Lattice Design (BOLD) method for Phase I clinical trials is extended to address an important challenge. It is widely understood that conventional Phase I trial designs are not consistently effective in determining safe and active dose levels. The US FDA launched the Project Optimus, aimed at reforming the paradigms of dose optimization and selection. We propose a backfill BOLD design (BF-BOLD) that centers on BOLD for dose-finding but also adds an activity evaluation for each patient. Our method for determining the optimal biological dose (OBD) first involves identifying the maximum tolerated dose (MTD) and then assessing activity rates among dose levels below the identified MTD. This approach is straight-forward and does not require complex statistical modeling. The results of the simulation indicate that performing dose-finding trials with backfilling can both enhance safety and activity assessment, thereby improving treatment sustainability while also preserving the potential for efficacy of the Recommended Phase II Dose (RP2D). We also demonstrate the applicability of the backfill design for reducing overdose rates, and as a more attractive alternative to small-scale dose expansion trials that follow dose escalation. Backfill designs are an important design approach for early phase trials.

## 1. Introduction

Phase I clinical trials aim to establish the maximum tolerable dose (MTD), defined as the dose with dose-limiting toxicity (DLT) rate closest to a specified target rate.^1^ The trials consist of three main components. The first component includes dose selection rules to determine dose level assignments. The second component involves deciding when to stop the trial. Note that given the potential for toxicity exposure for patients, Phase I trials generally are designed to parsimoniously enroll patients. A third component is a procedure to select a dose level as the MTD at the conclusion of the trial.^2^ From both a practical and ethical standpoint, it is essential to limit MTD identification errors in order to minimize the likelihood of subjecting patients to insufficiently therapeutic or overly toxic dosages in the next phase efficacy testing.^1^

Numerous Phase I trial designs have been developed to identify the MTD for single-agent studies, one of which is the Bayesian Ordered Lattice Design (BOLD) established by Wang and Tatsuoka (2026).^3^ The term “lattice” originates from the idea that the dose levels lie within a formal order structure, while the dose levels in single-drug trials can be perceived as a linearly ordered lattice.^4^ Figure S1 of the Supplementary Material depicts the lattice diagram that encompasses 5 dose levels, which also features both top and bottom states. The bottom state indicates that the MTD is less than the minimum dose. Conversely, the top state indicates that the MTD exceeds the maximum dose. Classification is associated with a specific state within the lattice, while a designated dose level signifies that the dose corresponds to the MTD. Relevant research on Bayesian sequential classification within lattice models includes works by Tatsuoka and Ferguson (2003),^5^ Tatsuoka (2014),^6^ and Tatsuoka, Chen, and Lu (2022).^7^

The BOLD design combines prior knowledge and order constraints with empirical toxicity data through a Bayesian framework. This approach facilitates adaptive dose selection, early trial termination, and the classification of the MTD in Phase I clinical trials.^3^ The primary characteristic of BOLD lies in its integration of posterior information and the implementation of order constraints at each stage of decision-making, facilitating the sharing of information across different doses. It represents an innovative and computationally fast method that showcases distinct conceptual and broad performance benefits in Bayesian dose-finding trials compared to well-established designs such as the Bayesian Optimal Interval Design (BOIN), Continual Reassessment Method (CRM), and the conventional 3+3 design.^3^ The performance of BOLD for Phase I clinical trials can be evaluated via simulations conducted with R (code available at https://github.com/hiddenmanna1996/BOLD).

In addition to the assessment of toxicity, the evaluation of *activity* is crucial in clinical trials. Activity refers to the capacity of an intervention, such as a cancer treatment drug, to achieve the intended positive outcome, such as target engagement, mechanistic response, or improved clinical outcome. The emergence of molecularly targeted agents and immunotherapies, characterized by toxicities that plateau in rate as dosage increases, has resulted in the formulation of the optimal biological dose concept, which takes into account both therapeutic activity and associated toxicity.^8^ In conventional practice, Phase I trials primarily concentrate on assessing the safety of the treatment using a limited sample size, and Phase II trials aim to evaluate activity by increasing the sample size at RP2D (recommended Phase II dose).

However, it has been recognized that the traditional approach for assigning the identified MTD as RP2D might not be sufficient, and it is widely acknowledged that standard Phase I trial designs can fail to consistently determine safe, active and sustainable doses.^9^ The US FDA has launched the Project Optimus, aimed at reforming the paradigms of dose optimization and selection. The more sophisticated notion of dose optimization aims to determine a dose level that maintains activity while ensuring the tolerability of adverse effects in practice. A new generation of dose-finding trials will identify the MTD while estimating activity at dose levels. When a trial can incorporate an evaluation of treatment activity within the same period necessary to assess toxicity, it is especially reasonable to take into account both toxicity and activity into account when determining the appropriate RP2D dose. This gives rise to the notion of an optimal biological dose (OBD) that balances these concerns, which may be lower than the MTD. Using a dose lower than MTD may enhance longer-term tolerability. Hence, OBD identification is an important practical objective for modern drug trials.

The approach to achieve this objective involves allocating patients to lower dose levels in order to evaluate activity during the dose-finding procedure, referred to as *backfilling*.^10^ Patients are enrolled in this manner to gain more information about lower dose level activity while waiting for dose toxicity responses in cohorts of patients assigned a current dose level. A motivation for collect more data this way is that the lower dose levels, being below the current dose level in dose-finding, are more likely to be safe relative to a target toxicity rate. The backfill strategy is particularly beneficial for OBD identification.^11^ A main objective of backfilling is to assess whether there is a “plateau” in the activity rates of a drug preceding the MTD. If such a plateau can be identified, it would be advantageous to instead select the lowest dose on this plateau rather than the MTD itself. This may enable finding a dose that preserves a similar level of activity while potentially reducing toxicity.

The aim of this study is thus to expand the application of the BOLD design, adapting it to address the challenge of dose optimization by means of backfilling. Backfill designs are compelling. Below, we show how backfill designs can improve MTD accuracy, reduce overdose rates especially when used in conjunction with existing overdose controls, can have high specificity for determining that activity rates for dose levels at or below MTD are below a given activity rate threshold, and are attractive alternatives to add-on dose expansion studies (Phase 1b).

## 2. Backfills in Phase I Clinical Trials

There are numerous methodologies for backfilling strategies in the early phases of clinical trials. Dehbi (2021) introduced an algorithm that involves randomly backfilling patients to dose levels that are lower than the currently administered dose, while employing Bayesian CRM as the framework for the toxicity trial to determine the MTD.^10^ Backfilling to the lower dose levels is stopped when they demonstrate inadequate activity in comparison to higher doses, beginning with the lowest dose. Although the toxicity data from backfilled patients is not incorporated into the toxicity trial, the activity data from all patients, including both backfilled and dose-finding participants, are utilized to establish the activity plateau for determining the optimal dose.

BF-BOIN (Backfilling incorporated into the BOIN design) has implemented a backfill strategy utilizing the BOIN algorithm in Phase I clinical trials.^12^ The doses available for back-filling are those that are lower than the currently administered dose, provided that at least one response has been observed at or below that level. If the DLT rate for a specific dose, along with the pooled DLT rate for that dose and its corresponding cover dose, exceeds the de-escalation threshold established by the BOIN design, that particular dose will be removed from consideration as a backfill candidate. A cohort of patients will be backfilled at the highest available dose. At the end of the trial, toxicity data from all doses, including those from both the toxicity trial and backfills, will be gathered to determine the MTD. See also for instance the backfill i3+3 design, an extension of the i3+3 framework specifically for backfilling.^13^

## 3. BF-BOLD: Integrating Backfilling into BOLD

The BOLD algorithm for identifying the MTD can be adapted for backfilling to conduct optimal biological dose finding. This approach, referred to as BF-BOLD, is geared towards identifying the safest dose that also has a “close enough” activity rate to MTD. The general premise is that, as in a standard Phase I trial, we collect toxicity data for each patient. We assume that these data are binary (a DLT has occurred or not). Additionally, for each patient, we assume a binary activity response. We continue to assume that dose selection and stopping decisions are driven by dose finding to identify the MTD, and this part of the proposed backfill design follows as in Wang and Tatsuoka (2026).^3^ MTD identification will also follow the same approach. We thus focus on how to backfill during the dose finding portion of a study and how to conduct OBD identification after an MTD has been selected. We also study the impact of backfilling on MTD and OBD accuracy, and overdose rates.

### Notation and Terminology

Terminology and notation of BF-BOLD follow as for BOLD,^3^ with added notation for activity data. Suppose we have *J >* 1 doses. Define DLT rate for dose *j* as *π*_*j*_, 1 ≤ *j* ≤ *J*. We assume the natural order constraints *π*_*j*_ ≤ *π*_*k*_ if *j < k*. We also define the *cover* dose for dose *j* to be dose *j* + 1, *j < J*, and the *anti-cover* to be dose *j* − 1, *j >* 1. Let *ϕ* be the target DLT rate. *N*_*max*_ is the maximum number of patients assigned through a dose finding rule in the trial before stopping is invoked, and does not include backfills; 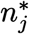 is the upper limit of patients at dose *j* before stopping of the trial is considered, including backfilling; *n*_*c*_ is the number of patients per cohort at a given stage for a dose-finding selected dose, where a *stage* is defined as the period of administration and observation for a DLT after a dose has been selected; *n*_*j*_ represents the total number of patients who have received dose level *j*, and *x*_*j*_ denotes the number of those patients with observed DLTs, with 0 ≤ *x*_*j*_ ≤ *n*_*j*_, including backfills. The activity outcome, represented by 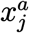 for dose level *j*, is also presumed to be binary and to follow a Binomial distribution characterized by the activity rate 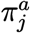. Additionally, *γ*_*j*_ represents the toxicity threshold for dose *j*, in that if the posterior probability of *π*_*j*_ *> ϕ* exceeds *γ*_*j*_, then the administration for all doses *k, j* ≤ *k*, will no longer be considered. Also, *τ* is a parameter indicating a target probability value used in dose selection.

### Prior and Conjugate Posterior Distributions

Given that that a DLT is a binary outcome, we employ Beta distributions, which are represented as *π*_*j*_ ∼ *Beta*(*α*_*j*_, *β*_*j*_) for dose level *j*. Importantly, Beta distributions have conjugate posteriors *π*_*j*_|*x*_*j*_, *n*_*j*_ ∼ *Beta*(*α*_*j*_ + *x*_*j*_, *β*_*j*_ + *n*_*j*_ − *x*_*j*_). In the case of trials assuming non-informative priors, we define the parameters of the prior distribution *α* and *β* by setting the prior mean equal to the target rate, while the prior effective sample size (PESS), *α* + *β*, is suggested to match the standard cohort size (e.g., *n*_*c*_ = 3) at a given stage of administration. For activity, it is assumed that 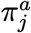 follows a uniform distribution of Beta(1,1), whereas the mean activity rate is 0.5, PESS = 2, and the activity rate spans from 0 to 1. As a result, the conjugate distribution of 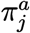 follows a Beta distribution 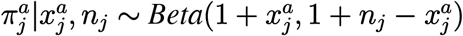 once the activity outcome 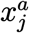 and sample size *n*_*j*_ have been observed with 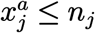.

### Toxicity Control

To control the dose toxicity on behalf of patient safety, we define CPAT to be the Conjugate Posterior Probability of the DLT rate being Above the Target, expressed as

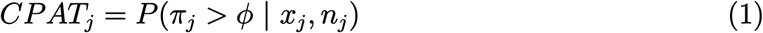

During experimentation, we will dynamically remove doses that are considered excessively toxic. Given a threshold *γ*_*j*_ *>* 0, a dose *j* is excessively toxic if *CPAT*_*j*_ *> γ*_*j*_. Note that if deemed overly toxic, dose *j* and all doses *k, j* ≤ *k*, are removed from further consideration. Furthermore, each dose can have its own threshold. The default value of *γ*_*j*_ is established at 0.95, with the exception of *γ*_1_, which is designated at 0.9 to enforce stricter toxicity control at the lowest dose.

### Backfill Implementation

The main factor in determining an appropriate backfilled dose is safety. It is crucial to avoid assigning patients a dose that lacks adequate evidence to verify its safety. Consequently, the dose for backfilling must be lower than the dose currently being administered in the dose-finding portion of the trial that focuses on identifying the MTD with methods from BOLD. In addition, backfill designs must reduce the likelihood of administering potentially inactive agents and/or sub-therapeutic doses to additional patients.^12^ We define *CPAT*^*a*^ as the Conjugate Probability of the Activity Rate being Above the Target, expressed in (2), where “Target” means the minimum acceptable activity rate represented by *ϕ*^*a*^.

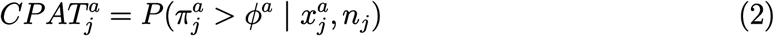

If the value of *CPAT*^*a*^ for dose *j* is below *γ*^*a*^ (with default value of 0.2), then dose *j* and all doses below *j* will be considered insufficiently active and not eligible for backfilling. Such an activity control is performed whenever backfill is employed at each stage of the trial. A dose that satisfies all of these criteria is considered an appropriate candidate for backfilling. In the proposed BF-BOLD design, patients are only allocated patients to the anti-cover of the dose being administered in the dose-finding part of the trial for backfill, provided it meets eligibility criteria. The size of the cohort for backfill may vary from 1 to 3, contingent upon availability and design.

### Order Constraints on Toxicity

Order constraints on CPAT values for toxicity are implemented after each stage using the Pool-Adjacent-Violators Algorithm (PAVA),^14^ an isotonic regression method that is appropriate for toxicity which is presumed to be non-decreasing with dose.^15,16^ PAVA ensures that the resulting estimates exhibit monotonicity and it is applied in a “local” manner, in the sense that only the CPATs of the current dose administered in the MTD-finding trial as well as its cover and anti-cover doses are included. A corresponding constrained *CPAT*_*j*_ value following PAVA is referred to as the PAVA-adjusted Posterior Probability of the DLT rate being Above the Target (*PPAT*_*j*_), which are the basis for dose selection in dose-finding trial.

### Dose Selection for Dose-finding

Dose selection in MTD-finding trial is restricted to either escalation, de-escalation, or maintaining the current dose being administered. The proposed selection criterion involves identifying the dose for which the PPAT is nearest to *τ*, the PPAT threshold parameter, among the current dose and its cover/anti-cover doses, where 0 *< τ* ≤ 0.5. The default value of *τ* is 0.5, and a PPAT value of 0.5 signifies the highest level of uncertainty regarding the safety of that dose level (akin to a coin flip). The *τ* value can be adjusted slightly downwards to serve as an overdose control parameter.^3^ Note that doing so encourages the most attractive dose to have uncertainty as to its toxicity but also to hedge towards safety. If we denote this set of dose levels as *J* ^∗^, then dose *j*^∗^ ∈ *J* ^∗^ is selected if it minimizes (3):

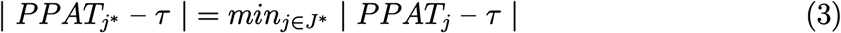

### Stopping Rule

The trial utilizing BF-BOLD is halted once one of the following conditions is met: 1) The minimum dose (i.e., dose 1) is excessively toxic, as indicated by *CPAT*_1_ *> γ*_1_, suggesting that the MTD is lower than this minimum dose level; 2) The overall count of patients who are allocated for dose-finding (not including the patients for backfilling) has reached a designated figure (e.g., *N*_*max*_ = 21 or 30); 3) The overall number of patients at the currently administered dose *j*, including those observed from dose-finding selected cohorts and participants in the backfill process, has reached a specified maximum limit 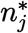, and dose *j* is the next stage selection in the dosefinding trial. In examples below, the value of 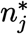 is set to be 12 for *j >*1, and 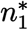 is set to be 15 in order to enhance classification to the bottom state, similar to the BOLD design.

### MTD Identification

The MTD is determined upon the stopping of the trial. MTD identification relies on isotonically-regressed posterior means of DLT rates. As noted, if the lowest dose is found to be excessively toxic (*CPAT*_1_ *> γ*_1_), all doses are considered too toxic and no MTD is selected. Otherwise, we implement PAVA “locally” on the conjugate posterior means of the final selected dose in dosefinding trial, along with its cover and/or anti-cover doses (if applicable), and regard these as the potential candidates. The candidates for MTD are required to have trial data. Let *K*^∗^ be the group of candidates, and 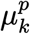 be the PAVA-adjusted posterior mean for dose *k*. The dose *k* ∈ *K*^∗^ with smallest PAVA-adjusted posterior mean difference with target DLT rate in (4) is selected as MTD.

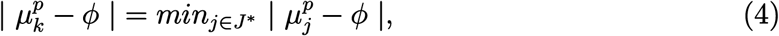

### OBD Identification

The optimal dose is also identified following the completion of the BOLD-based dose finding portion of the trial. The OBD is determined by the PAVA-adjusted posterior means of activity rates for doses that do not exceed the identified MTD (*M*). Local PAVA (for the dose of interest and its anti-cover and cover dose if applicable) will be applied. There are three thresholds for OBD. First, a reduction (trade-off) from the identified MTD for the PAVA-adjusted posterior mean activity rate is accepted for OBD, where the default trade-off value is set at 10% (or 5%, for example, user choice). Second, the PAVA-adjusted posterior mean activity rate of the OBD must be at least *ϕ*^*a*^, the minimum acceptable activity rate. Third, OBD cannot exceed *M*. The lowest dose which does not exceed *M* and whose PAVA-adjusted posterior mean activity rate is both at least *ϕ*^*a*^ and within the trade-off of the estimated activity rate of *M* will be identified as OBD. OBD is not determined if the minimum dose is considered to be excessively toxic.

### Flowchart of BF-BOLD

The flowchart illustrating the backfill integration with BOLD is displayed in Figure 1. A summarized description of the flowchart is as follows:

1. Specify the target DLT rate *ϕ*, the minimum acceptable activity rate *ϕ*^*a*^, activity threshold trade-off value, overdose control *τ* parameter, toxicity threshold *γ* (or *γ*_*j*_ for each dose *j*), *N*_*max*_ for dose-finding (excluding the patients designated for backfill), and 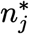 for each dose *j*.
2. Specify the prior information at dose level, such as prior mean DLT rates (default values = *ϕ*), prior distribution of activity rate (default is Beta(1,1)), and PESS values.
3. Determine the dose finding selection using BOLD, while starting with the lowest dose if the trial is at the initial stage (see Wang and Tatsuoka (2026) for more details).
4. Administer the selected dose to a cohort of patients (default *n*_*c*_ = 3) to observe data on both toxicity and activity.
5. Monitor toxicity at dose level, and stop the trial with no MTD or OBD identified if the lowest dose is overly toxic.
6. Monitor whether the maximum sample size limit for dose finding (e.g., *N*_*max*_ = 21 or 30) has been attained. If this is the case, stop the trial and go to Step 13.
7. Observe whether the cumulative count of treated patients, which includes the patients in both dose finding and backfill, at the current dose being administered for dose finding attains its maximum (e.g., 12 or 15) when the decision is to maintain the current dose. If this is the case, stop the trial and go to Step 13.
8. Update *CPAT*^*a*^ values, which indicate the posterior probability of a dose being active, and assess whether any value of the doses that are lower than the current non-backfill dose level is at least *γ*^*a*^ (with a default value of 0.2) to determine its eligibility for backfill. If none is eligible, go to Step 12.
9. Determine the backfill dose (i.e., the anti-cover of the currently administered dose for dose finding) and assign patient(s) for up to the specified cohort size for backfill.
10. Observe both DLTs and dose activity of backfilled patients.
11. Update PPATs of toxicity locally around the current dose administered for dose finding by incorporating backfill data for posterior updating and dose selection.
12. Repeat Steps 3 to 11 until the trial stops.
13. Once stopping occurs, identify the MTD by considering PAVA-adjusted posterior mean DLT rate and OBD by PAVA-adjusted posterior mean activity rate, except when the lowest dose exhibits excessive toxicity, leading to the conclusion that there is no MTD or OBD.

**Figure 1.**
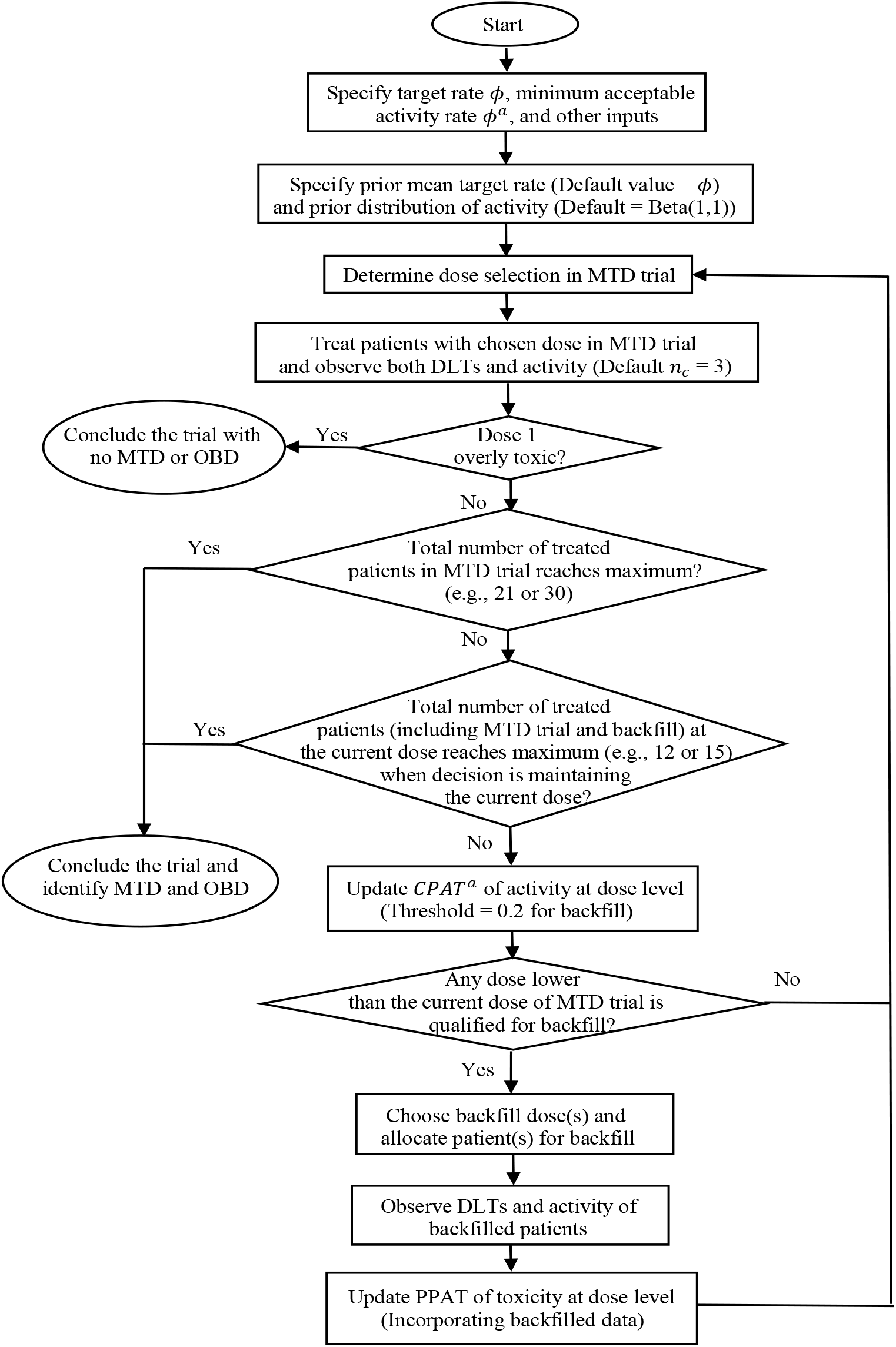
Flowchart of backfilling incorporated in BOLD design (BF-BOLD).

## 4. Simulation

### Demonstration of BF-BOLD

A demonstration of BF-BOLD design for single-drug trials is be presented by simulation (using R 4.4.3, code available at https://github.com/gxw174/BF-BOLD). Please refer to the notation and terminology defined above, and see Wang and Tatsuoka (2026) for the reasoning behind the design input selections. We assume that *J* = 5, *n*_*c*_ = 3, *ϕ* = 0.25, and *ϕ*^*a*^ = 0.5. For the dose-finding part of the trial (non-backfilling), *N*_*max*_ = 30 patients, 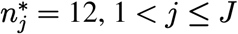, which includes backfills, and 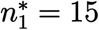. The true MTD and true OBD are dose level 4 and 2, respectively. The prior mean DLT rate is assumed to be equivalent to *ϕ* for all dose levels, and the prior distribution of activity rate is assumed to be Beta(1,1). The PESS values are set at 3. The Bayesian toxicity threshold, based on the CPAT criterion for identifying excessively toxic doses, is established at *γ*_*j*_ = 0.95 for *j >* 1 and *γ*_1_ = 0.9. The true DLT rates in the simulation are assumed to be 0.1, 0.12, 0.15, 0.25, and 0.4 for doses 1 to 5, respectively. The true activity rates are 0.55, 0.7, 0.7, 0.7, and 0.7, indicating that the activity stabilizes from dose level 2 onward. At each stage, 1 to 3 patients are randomly backfilled. Finally, a 10% trade-off of activity rate is utilized for the OBD identification.

The proposed procedure, involving dose escalations and backfilling at each stage, is illustrated in Figure 2. The trial comes to an end at the tenth stage when the total sample size for allocated for dose-finding reaches *N*_*max*_ = 30. Dose 4 is accurately recognized as the MTD, as its PAVA-adjusted posterior mean toxicity rate is closer to *ϕ* than for dose 5, the final administered dose, with corresponding values of 0.25 and 0.292 respectively. Dose 2 is correctly identified as the OBD, as the locally PAVA-adjusted posterior mean activity rates of dose 2 is within a 10% margin of the corresponding value for the identified MTD (dose 4) as well as being greater than *ϕ*^*a*^. These outcomes indicate that the proposed backfilling strategy can correctly identify the MTD and OBD.

**Figure 2.**
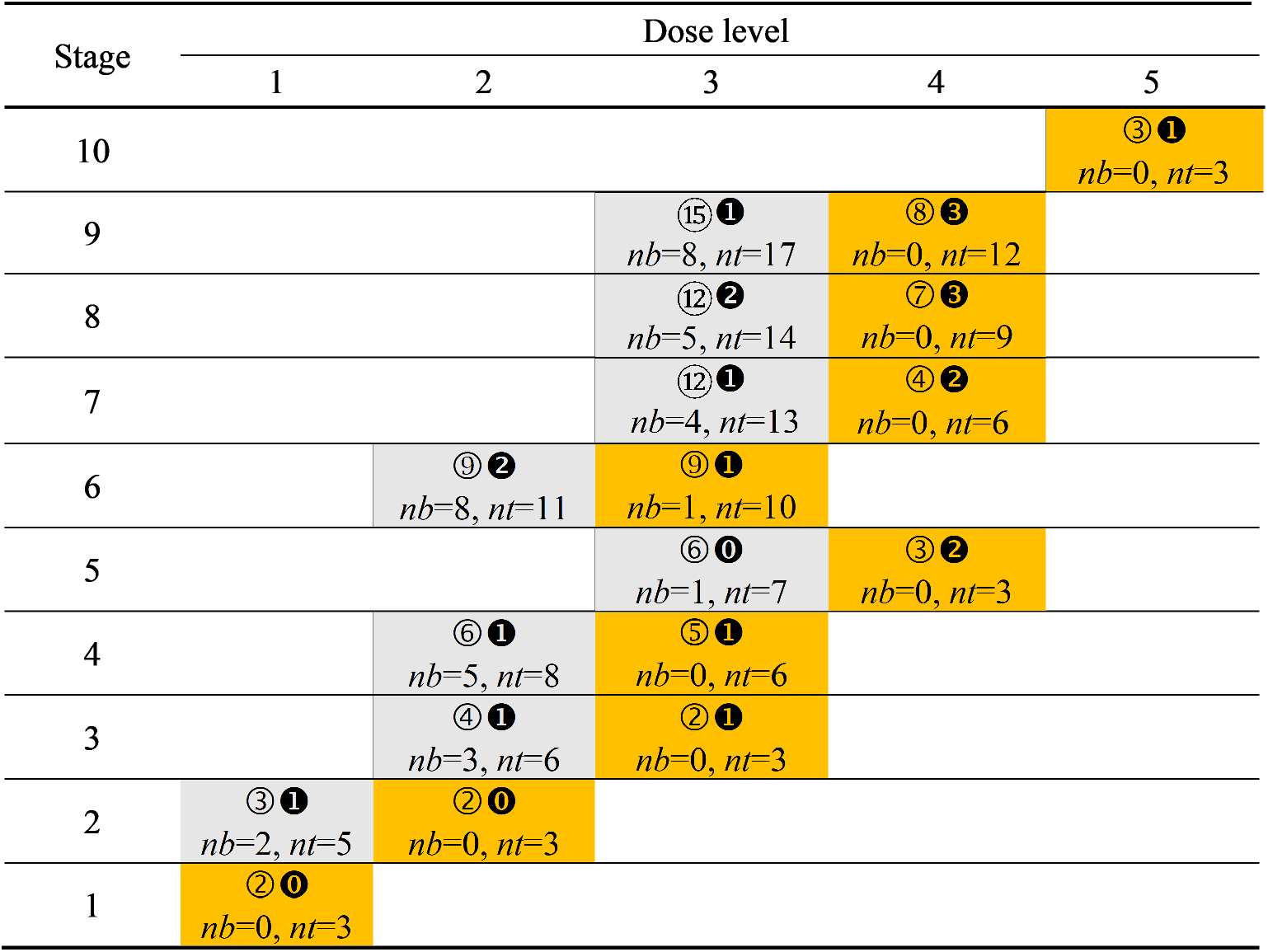
Diagram of the demonstration of dose selections and backfills at *ϕ* = 0.25, *ϕ*^*a*^ = 0.5, true MTD at level 4, true OBD at dose 2, and *N*_*max*_ = 30. The true DLT rates are presumed to be 0.1, 0.12, 0.15, 0.25, and 0.4 for doses 1 to 5, respectively. The true activity rates are 0.55, 0.7, 0.7, 0.7, and 0.7, indicating an activity plateau commencing from dose 2. Orange indicates dose finding, while gray signifies backfilling. Numbers displayed on a black background indicate accumulated DLTs, while numbers on a clear background signify accumulated activity counts. The value of *nt* indicates the sample size (including for dose finding and backfills) at dose level, and *nb* indicates the accumulated number of backfilled patients. Dose 4 is accurately recognized as the MTD at the 10th stage. Dose 2 is correctly identified as the OBD, as it signifies the minimum dose at which the local PAVA-adjusted posterior mean activity rates are within a 10% margin of the identified MTD (dose 4) and greater than *ϕ*^*a*^.

### Performance Analysis of BF-BOLD

The performance of BF-BOLD can be assessed through a range of simulations. Consider first a design consisting of 5 dose levels, where the possible MTD identifications encompass “*<*1”, 1, 2, 3, 4, 5, and “*>*5”, where “*<*1” indicates that all doses are too toxic, and “*>*5” signifies that all doses have DLT rates that fall below the target. Simulations are conducted using random scenarios that incorporate both true DLT and true activity rates. The lower and upper limit for DLT rates are set at 0.01 and 0.7 respectively. We assume that the difference between the true DLT rate for the cover dose of MTD and that of MTD, i.e., upper *δ*, is either 0.1, 0.15, 0.2 or 0.25, whereas the true DLT rate for the anti-cover dose of MTD is at least 0.05 lower than that of the MTD. All other dose-level DLT rates are randomly generated in an ascending order in a range between 0.01 and 0.7. For a specified upper *δ*, 50 random scenarios are created for each MTD level, with a total of 350 random scenarios. For each of the 350 random scenarios, 100 simulations of response sequences are generated.

For activity rates, OBD is assumed to be zero, one or two levels below the true MTD. We refer to the difference in dose levels between true MTD and true OBD as the *gap*. Hence, we assume gap = 0, 1 or 2. Let the true OBD be denoted as dose *d*^*a*^, so the true activity rate of dose *d*^*a*^ is established at 0.7. It is also assumed that a plateau of activity rate begins from dose *d*^*a*^, and the true activity rate of the anti-cover of dose *d*^*a*^ (i.e., dose *d*^*a*^ − 1 if it is present) is determined to be 0.7 minus the value of lower *δ*^*a*^, with the default value of lower *δ*^*a*^ set at 0.15. The true activity rates of the doses less than *d*^*a*^ − 1, as they exist, are randomly generated in an ascending order with a lower limit of 0.1 and an upper limit of 0.55.

Table 1 presents the simulation results of BF-BOLD design regarding the identification accuracies, efficiencies, overdose rates, underestimation rates, and overestimation rates with a 5-dose lattice at *ϕ* = 0.25, *ϕ*^*a*^ = 0.5, and *N*_*max*_ (for MTD dose finding) = 21, with both upper *δ* for random scenarios of true DLT rates and lower *δ*^*a*^ for random scenarios of true activity rates set at 0.15. Average accuracy rates of MTD and OBD identification arising from simulations, as well as the standard deviations, are reported across various scenarios. The accuracies of OBD identification are presented in three ways: the overall accuracy, the accuracy in scenarios where the identified MTD corresponds with the true MTD, and the accuracy when the identified MTD surpasses the true MTD. This allows for understanding the impact of overestimation of the MTD on OBD identification accuracy. The average number of patients treated and corresponding standard deviation across scenarios is presented, both overall and among backfilled patients. The overdose rate is the proportion of patients treated with doses higher than the true MTD, for both overall and backfilled patients. The underestimation rate is the proportion of trials with identified MTD/OBD being lower than the true MTD/OBD for a given scenario, and overestimation rate is the proportion of trials with identified MTD/OBD being higher than the true MTD/OBD for a given scenario. An activity plateau is assumed to be present, and a random number of backfilled patients (1 to 3) receive the anti-cover dose corresponding to the current dose being administered for dose finding. A 10% activity rate trade-off is utilized for the identification of OBD. Table S1 in the Supplementary Material illustrates the result for *N*_*max*_ set at 30. Tables S2 and S3 illustrate the result with lower *δ*^*a*^ for random scenarios of true activity rates set at 0.2, reflecting greater discrimination at the activity rate plateau.

**Table 1:** Summary of identification accuracy, number of treated patients, overdose rate, underes-timation rate, and overestimation rate of BF-BOLD design with 5-dose lattice at *ϕ* = 0.25, *ϕ*^*a*^ = 0.5, true activity rate of OBD = 0.7, upper *δ* (for DLT rates) = 0.15, lower *δ*^*a*^ (for activity rates) = 0.15, and *N*_*max*_ = 21, utilizing random scenarios of true DLT rates and activity rates, while OBD being zero, one or two levels below the true MTD, so that gap = 0, 1, 2. A random number of backfilled patients (1 to 3) receive the anti-cover dose corresponding to the current dose in the dose-finding trial. A 10% activity rate trade-off is utilized for the identification of OBD.

| Gap<br>(MTD<br>minus<br>OBD) | MTD<br>level | Accuracy of identification (SD) |  |  |  | Number of treated patients<br>(SD) |  | Overdose rate |  | Underestimation<br>rate |  | Overestimation<br>rate |  |
| --- | --- | --- | --- | --- | --- | --- | --- | --- | --- | --- | --- | --- | --- |
|  |  | MTD | OBD |  |  | Overall | Backfill<br>only | Overall | Backfill<br>only | MTD | OBD | MTD | OBD |
|  |  |  | Overall | When<br>identified<br>MTD = true<br>MTD | When<br>identified<br>MTD > true<br>MTD |  |  |  |  |  |  |  |  |
| 0 | <1 | 54.82 (14.00) | -- | -- | -- | 14.20 (2.16) | 1.43 (0.60) | 0.19 | 0.15 | -- | -- | 0.45 | -- |
|  | 1 | 66.02 (4.75) | 77.56 (4.39) | 94.79 (2.44) | 66.61 (11.06) | 21.06 (0.73) | 3.93 (0.36) | 0.37 | 0.21 | 0.12 | 0.15 | 0.22 | 0.07 |
|  | 2 | 56.42 (8.19) | 44.16 (7.58) | 61.09 (7.24) | 41.35 (9.55) | 26.20 (0.82) | 7.09 (0.76) | 0.23 | 0.08 | 0.20 | 0.49 | 0.23 | 0.07 |
|  | 3 | 48.18 (8.47) | 37.14 (7.51) | 59.05 (8.46) | 37.44 (11.02) | 26.91 (1.06) | 6.70 (0.88) | 0.16 | 0.04 | 0.28 | 0.56 | 0.23 | 0.07 |
|  | 4 | 43.08 (7.34) | 29.58 (6.88) | 51.47 (8.88) | 37.24 (9.73) | 26.53 (1.16) | 5.88 (1.13) | 0.09 | 0.00 | 0.37 | 0.65 | 0.20 | 0.06 |
|  | 5 | 51.46 (10.70) | 27.56 (6.33) | 53.88 (7.68) | -- | 25.84 (1.33) | 5.00 (1.29) | -- | -- | 0.49 | 0.72 | -- | -- |
|  | >5 | 78.94 (10.60) | 32.32 (8.48) | 41.06 (10.01) | -- | 25.61 (1.21) | 4.66 (1.22) | -- | -- | 0.21 | 0.68 | -- | -- |
|  | **Avg.** | **56.99 (9.15)** | **41.39 (6.86)** | **60.22 (7.45)** | **45.66 (10.34)** | **23.77 (1.21)** | **4.95 (0.89)** | **0.21** | **0.09** | **0.28** | **0.54** | **0.27** | **0.07** |
| 1 | <1 | 54.82 (14.00) | -- | -- | -- | 14.20 (2.16) | 1.43 (0.60) | 0.19 | 0.15 | -- | -- | 0.45 | -- |
|  | 1 | 66.02 (4.75) | -- | -- | -- | 21.06 (0.73) | 3.93 (0.36) | 0.37 | 0.21 | 0.12 | -- | 0.22 | -- |
|  | 2 | 57.50 (9.18) | 69.44 (5.18) | 67.24 (6.80) | 55.36 (9.99) | 26.65 (0.80) | 7.58 (0.75) | 0.23 | 0.06 | 0.20 | 0.03 | 0.22 | 0.27 |
|  | 3 | 48.24 (8.19) | 42.46 (5.28) | 43.51 (7.74) | 28.93 (9.27) | 28.43 (0.98) | 8.27 (0.73) | 0.15 | 0.03 | 0.28 | 0.35 | 0.24 | 0.22 |
|  | 4 | 42.72 (7.78) | 38.06 (5.58) | 40.40 (7.46) | 29.17 (12.04) | 27.88 (0.80) | 7.23 (0.72) | 0.08 | 0.00 | 0.37 | 0.42 | 0.20 | 0.20 |
|  | 5 | 51.76 (11.07) | 34.90 (6.13) | 35.58 (8.27) | -- | 27.11 (1.13) | 6.27 (1.07) | -- | -- | 0.48 | 0.49 | -- | 0.16 |
|  | >5 | 78.64 (10.20) | 43.28 (7.93) | 54.92 (6.33) | -- | 26.59 (1.06) | 5.64 (1.04) | -- | -- | 0.21 | 0.57 | -- | -- |
|  | **Avg.** | **57.10 (9.31)** | **45.63 (6.02)** | **48.33 (7.32)** | **37.82 (10.43)** | **24.56 (1.09)** | **5.77 (0.75)** | **0.20** | **0.09** | **0.28** | **0.37** | **0.27** | **0.22** |
| 2 | <1 | 54.82 (14.00) | -- | -- | -- | 14.20 (2.16) | 1.43 (0.60) | 0.19 | 0.15 | -- | -- | 0.45 | -- |
|  | 1 | 66.02 (4.75) | -- | -- | -- | 21.06 (0.73) | 3.93 (0.36) | 0.37 | 0.21 | 0.12 | -- | 0.22 | -- |
|  | 2 | 57.50 (9.18) | -- | -- | -- | 26.65 (0.80) | 7.58 (0.75) | 0.23 | 0.06 | 0.20 | -- | 0.22 | -- |
|  | 3 | 46.62 (8.67) | 56.94 (5.89) | 52.49 (8.47) | 49.82 (11.06) | 28.64 (1.03) | 8.53 (0.71) | 0.15 | 0.03 | 0.30 | 0.01 | 0.24 | 0.42 |
|  | 4 | 43.64 (8.28) | 35.18 (5.83) | 29.11 (7.12) | 27.05 (10.58) | 29.23 (0.64) | 8.61 (0.51) | 0.08 | 0.00 | 0.37 | 0.32 | 0.20 | 0.33 |
|  | 5 | 51.08 (10.60) | 34.50 (4.82) | 28.07 (5.85) | -- | 28.36 (0.86) | 7.54 (0.76) | -- | -- | 0.49 | 0.35 | -- | 0.30 |
|  | >5 | 78.08 (11.36) | 36.76 (4.83) | 36.24 (4.66) | -- | 27.79 (0.90) | 6.85 (0.87) | -- | -- | 0.22 | 0.39 | -- | 0.24 |
|  | **Avg.** | **56.82 (9.55)** | **40.85 (5.34)** | **36.48 (6.52)** | **38.44 (10.82)** | **25.13 (1.02)** | **6.35 (0.65)** | **0.20** | **0.09** | **0.28** | **0.27** | **0.27** | **0.32** |

These tables indicate that the accuracy levels in OBD identification fluctuate depending on the gap between MTD and OBD, whereas the accuracy of MTD remains nearly unchanged irrespective of the gap. Note that it is hypothetically possible for backfill dose selection and hence dose finding to be affected by the gap size. The means and standard deviations of the number of patients receiving treatment show a slight increase as the gap widens. The overdose rate, as well as the underestimation and overestimation rate with respect to MTD, remain unchanged irrespective of the gap. The underestimation and overestimation rates for OBD diminish and escalate, respectively, as the gap expands.

### Overdose Control Analysis of BF-BOLD

Overdose control is also explored by adjusting the PPAT threshold parameter (*τ*) which can lead to a reduction in the overdosing rate.^3^ A summary is provided regarding the accuracy, efficiency, overdose rate, underestimation rate, and overestimation rate of BF-BOLD at a 5-dose model with *N*_*max*_ = 21 and 30, as shown in Table S4 and S5, respectively, while the value of *τ* is set to be 0.45 (0.5 is the default value). All the design characteristics, parameter settings, and analysis procedures are identical to those in Tables 1 and S1. The results show that the over-dosing rates with *τ* = 0.45 are significantly reduced in comparison to *τ* = 0.5. Additionally, it demonstrates that the overall MTD accuracy rates experience only a slight decline with a lower *τ* as a trade-off, due to less accuracy for higher dose levels.

### Specificity in OBD Identification for BF-BOLD

We additionally assess the performance of BF-BOLD when none of the true activity rates across the doses exceed *ϕ*^*a*^, the minimum acceptable activity rate, which is set to be 0.5 in our examples. This reflects the absence of an OBD. The correct OBD classification is thus to identify that no OBD exists. An example summary is presented of the accuracy, efficiency, overdose rate, underestimation rate, and overestimation rate of a 5-dose model in Table S6. The true activity rate of MTD is assumed at 0.3 and 0.4, a value that is lower than *ϕ*^*a*^ = 0.5. The accuracy rate of OBD identification refers to the likelihood of no OBD identification, also known as specificity. The table indicates that the overall specificity values for OBD with *N*_*max*_ = 21 are 0.93 and 0.79 when the true activity rate of MTD is 0.3 and 0.4, respectively, and they are 0.95 and 0.82 when *N*_*max*_ = 30. These show high specificity, which has important practical ramifications for Phase 1 trials in terms of the ability to inform potential lack of efficacy while establishing safety and MTD identification. Lack of sufficient activity rate could be an important factor in deciding whether or not to proceed to a next phase of testing.

### Comparative Analysis of BF-BOLD Backfill Strategies

We also examine the effects of various backfill approaches in clinical trials employing the BF-BOLD design, including the original BOLD design without backfill implementation but with activity observed, BF-BOLD with randomized 1 to 3 backfilled patients, and BF-BOLD with only one patient assigned for backfilling.

Table 2 provides a summary of the average accuracy (MTD and OBD), patient count (both overall and backfilled) and overdose rate (overall and backfilled) for a 5-dose model, stratified by *N*_*max*_ for dose-finding (21 and 30), gap between true MTD and true OBD (0, 1, and 2), and the number of backfilled patients per stage (0, 1, and randomized from 1 to 3). Note that the zero number of backfilled patients per stage corresponds to the BOLD design with no backfilling. All other assumptions remain consistent with those outlined in Table 1. The table indicates that the accuracies of MTD and OBD improve slowly as the number of backfilled patients increases, at the cost of a higher overall patient count. Additionally, the overdose rate declines as the number of backfilled patients increases, supporting the notion that backfilling enhances safety. Note that if only one patient is backfilled per stage, there is only a small incremental improvement in MTD and OBD accuracy, without too much increase in patient numbers or overdosing. Backfilling is more fruitful in improving MTD and OBD accuracy with 1-3 patients per stage (e.g. taking in as many patients as can be enrolled while waiting for the cohort assigned in dose-finding to complete their observation periods).

**Table 2:** Comparative analysis of accuracy, efficiency, and overdose rate among different backfilling strategies of BF-BOLD for 5-dose lattice, stratified by *N*_*max*_, gap between true MTD and true OBD, and the number of backfilled patients per stage. The notation of {1,2,3} signifies the random selection of 1 to 3 patients, while the BF-BOLD with no backfilled patients represents the original BOLD design without backfill implementation but with activity observed. All the other design characteristics and parameter settings are identical to those in Table 1.

| N<sub>max</sub><br>for<br>MTD<br>trial | Gap<br>(MTD<br>minus<br>OBD) | Number of<br>backfilled<br>patients<br>per stage | Accuracy of identification (SD) |  |  |  | Number of treated patients<br>(SD) |  | Overdose rate |  |
| --- | --- | --- | --- | --- | --- | --- | --- | --- | --- | --- |
|  |  |  | MTD | OBD |  |  | Overall | Backfill<br>only | Overall | Backfill<br>only |
|  |  |  |  | Overall | When<br>identified<br>MTD = true<br>MTD | When<br>identified<br>MTD > true<br>MTD |  |  |  |  |
| 21 | 0 | 0 | 54.51 (8.80) | 38.70 (6.44) | 57.52 (7.73) | 39.39 (8.86) | 19.24 (0.41) | -- | 0.25 | -- |
|  |  | 1 | 55.95 (8.89) | 40.46 (6.91) | 59.34 (7.44) | 41.19 (9.58) | 21.57 (0.76) | 2.46 (0.43) | 0.23 | 0.11 |
|  |  | {1,2,3} | 56.99 (9.15) | 41.39 (6.86) | 60.22 (7.45) | 45.66 (10.34) | 23.77 (1.21) | 4.95 (0.89) | 0.21 | 0.09 |
|  | 1 | 0 | 54.51 (8.63) | 40.39 (5.52) | 41.81 (6.67) | 33.41 (9.00) | 19.24 (0.41) | -- | 0.25 | -- |
|  |  | 1 | 55.78 (8.86) | 43.75 (6.21) | 45.35 (7.03) | 36.21 (9.85) | 21.98 (0.73) | 2.87 (0.38) | 0.23 | 0.11 |
|  |  | {1,2,3} | 57.10 (9.31) | 45.63 (6.02) | 48.33 (7.32) | 37.82 (10.43) | 24.56 (1.09) | 5.77 (0.75) | 0.20 | 0.09 |
|  | 2 | 0 | 54.30 (8.69) | 36.94 (5.04) | 33.12 (6.57) | 35.78 (8.77) | 19.24 (0.41) | -- | 0.25 | -- |
|  |  | 1 | 55.81 (8.85) | 38.05 (4.86) | 34.61 (6.73) | 38.96 (9.79) | 22.28 (0.67) | 3.17 (0.31) | 0.23 | 0.10 |
|  |  | {1,2,3} | 56.82 (9.55) | 40.85 (5.34) | 36.48 (6.52) | 38.44 (10.82) | 25.13 (1.02) | 6.35 (0.65) | 0.20 | 0.09 |
| 30 | 0 | 0 | 60.17 (7.86) | 43.12 (6.92) | 59.91 (7.43) | 40.17 (9.17) | 23.26 (0.69) | -- | 0.29 | -- |
|  |  | 1 | 60.73 (8.32) | 44.80 (7.13) | 62.11 (7.35) | 43.61 (10.99) | 26.34 (1.06) | 3.62 (0.56) | 0.26 | 0.13 |
|  |  | {1,2,3} | 62.01 (9.05) | 46.20 (7.28) | 64.16 (6.74) | 46.10 (11.37) | 29.02 (1.51) | 7.05 (1.12) | 0.23 | 0.11 |
|  | 1 | 0 | 59.73 (7.96) | 43.46 (5.45) | 43.44 (7.01) | 34.14 (8.31) | 23.25 (0.70) | -- | 0.29 | -- |
|  |  | 1 | 60.77 (8.79) | 47.57 (5.48) | 48.40 (6.65) | 37.01 (9.53) | 26.95 (0.95) | 4.22 (0.44) | 0.26 | 0.13 |
|  |  | {1,2,3} | 62.16 (9.25) | 49.84 (5.67) | 51.40 (6.31) | 37.05 (10.21) | 30.17 (1.28) | 8.24 (0.84) | 0.23 | 0.11 |
|  | 2 | 0 | 59.95 (7.93) | 35.76 (4.57) | 32.99 (5.59) | 36.67 (10.69) | 23.25 (0.70) | -- | 0.29 | -- |
|  |  | 1 | 60.81 (8.59) | 39.13 (5.62) | 35.72 (6.71) | 38.25 (10.63) | 27.30 (0.91) | 4.62 (0.37) | 0.26 | 0.13 |
|  |  | {1,2,3} | 61.91 (9.18) | 42.31 (5.25) | 39.01 (6.55) | 39.71 (10.39) | 30.92 (1.25) | 9.02 (0.75) | 0.22 | 0.11 |

### Comparative Analysis of BF-BOLD versus BF-BOIN

Comparative analyses are also conducted. Given that BOIN was chosen as the comparator for the BOLD design due to their inherent similarities,^3^ we use BF-BOIN as the reference for BF-BOLD performance. Recall that by default BF-BOLD is assumed to backfill a random number of patients with the anti-cover dose of the current dose used in dose-finding. This approach makes the two designs comparable, as BF-BOIN also assumes a time frame for the toxicity data from the dose-finding trial to be available before any dose decisions can be made, similarly leading to randomness in the cohort size of backfill patients.

Figures 3 presents the bar charts that compare the accuracy of MTD identification between BF-BOLD and BF-BOIN, stratified by *N*_*max*_ and upper *δ* value for dose-finding trial, for a 5-dose model. This comparison employs random scenarios of true DLT rates and activity rates for simulations, while the OBD is presumed to correspond to MTD (i.e., gap = 0) and a plateau of activity is present. The values of *ϕ, ϕ*^*a*^, and true activity rate of OBD are established at 0.25, 0.5 and 0.7 respectively as before, and the lower *δ*^*a*^ for activity is set to be 0.15. These figures show that BF-BOLD surpasses BF-BOIN in accurately identifying the MTD across all dose levels and upper *δ* values for *N*_*max*_ being both 21 and 30, with the exception when all doses are too toxic. Recall that the activity rates of OBD are not provided by BF-BOIN. The bar charts illustrating the efficiency, denoted by patient count and standard deviation, is displayed in Figure S2. BF-BOIN generally has higher average patient counts overall, although they are lower for lower doses, and higher for higher doses. The comparisons between BF-BOLD and BF-BOIN for gap = 1 and 2, which do not have bar charts displayed, exhibit a similar pattern.

**Figure 3.**
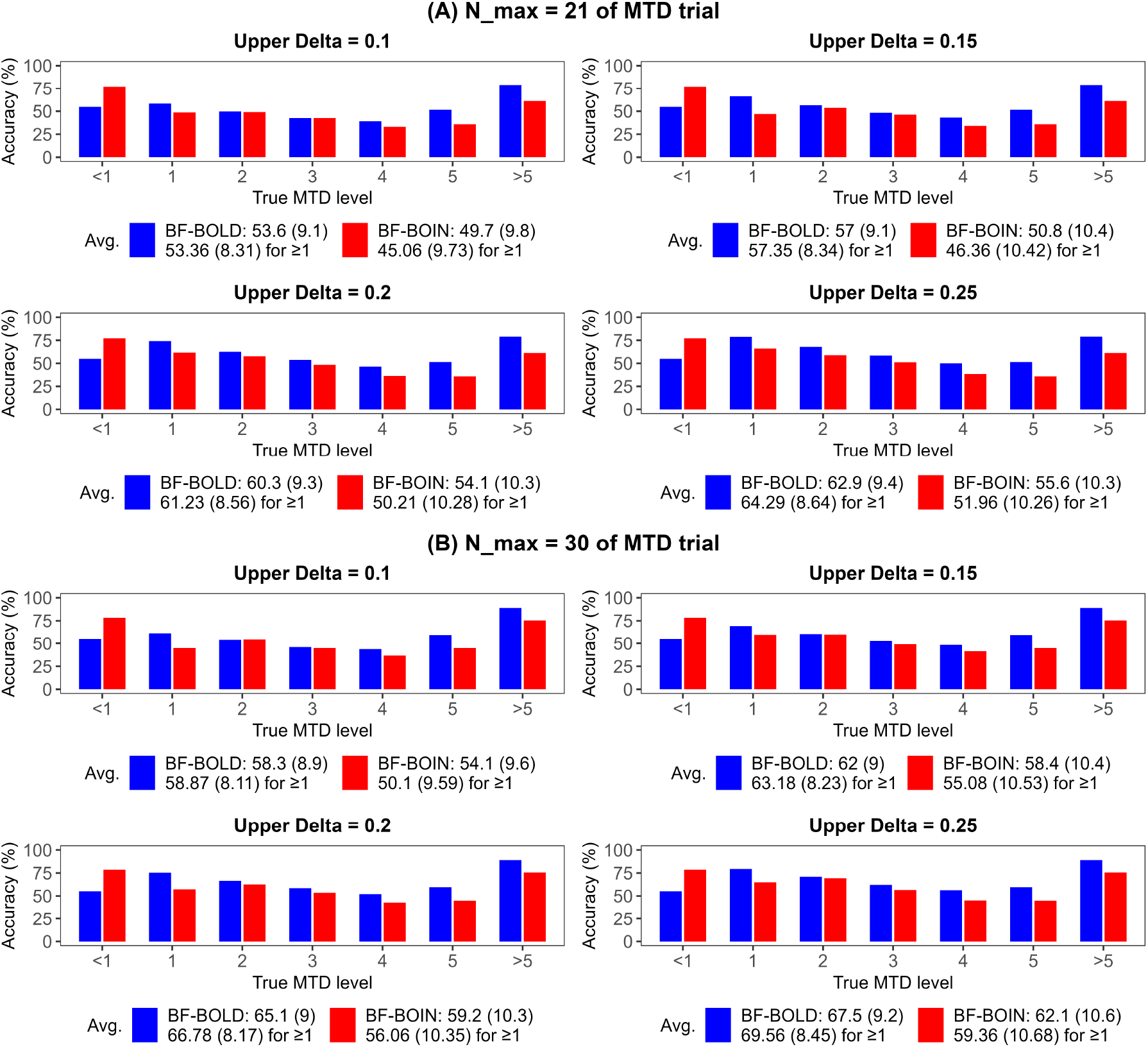
Bar charts illustrating comparative analysis of MTD accuracy (with standard deviation) between BF-BOLD and BF-BOIN for 5-dose lattice, stratified by *N*_*max*_ and upper *δ* of DLT rate, while OBD is assumed to be zero level below MTD (gap = 0) and a plateau of activity is present. All the other design characteristics and parameter settings are identical to those in Table 1. The legend denotes the average for all the 7 MTD levels (1st row) and for the MTD levels that are ≥ 1 (2nd row).

In addition to the 5-dose model, the performance summary of BF-BOLD for a 3-dose model in comparison to BF-BOIN, categorized by gap, is displayed in Tables S7 and S8 for *N*_*max*_ values of 21 and 30, respectively, with upper *δ* = 0.20. These tables also demonstrate MTD accuracy improvements for BF-BOLD when compared to BF-BOIN, except when all doses are excessively toxic, as well as smaller standard deviation in accuracy rates across randomly generated scenarios.

### Comparative Analysis of BF-BOLD versus BOLD-1b (Phase 1a/1b Dose Finding plus Expansion)

Here, we consider when the activity marker coincides with a clinical efficacy outcome, such as whether or not a major pathological response is achieved in cancer trials. This can happen for instance in a neoadjuvant trial, where tissue obtained at time of surgery can be analyzed for disease progression. In that case, a comparison can be conducted between using BF-BOLD to assess safety and efficacy versus conducting a phase 1a/1b study using BOLD in phase 1a dose-finding and then using the identified MTD as the single dose in a subsequent dose-expansion phase 1b study. Note that in identifying the OBD, we assume here that BF-BOLD can rely on the dose-expansion efficacy outcome as the activity marker. To evaluate the performance of BF-BOLD in comparison to such a combination Phase 1a/1b expansion design, we introduce the BOLD-1b design, which serves as an extension of the original BOLD design. BOLD-1b is distinguished from BOLD by two key features: 1) In addition to DLTs, activity counts are monitored at each stage of the trial; 2) At the end of the BOLD-designed Phase 1a dose expansion trial, we administer the resultant dose identified as MTD to a certain number of patients (e.g., 10). After the dose expansion phase, the methodology in BOLD-1b for updating the identified MTD and OBD remains consistent with that used in BF-BOLD, using the aggregated data.

Table 3 provides a summary of accuracy (MTD and OBD) and patient count at dose level for BF-BOLD versus BOLD-1b for a 5-dose model categorized by gap. Table S9 presents the summary of the corresponding overdose rates, and the underestimation and overestimation rates (MTD and OBD). Other assumptions, such as *ϕ, ϕ*^*a*^, true activity rate of OBD, upper *δ* and lower *δ*, remain consistent with those presented in Table 1. In the case of BF-BOLD, we randomize between 1 to 3 patients for backfilling at each stage, whereas BOLD-1b does not implement any backfilling. The values of *N*_*max*_ for the dose-finding part of the trial are established at 24 for BF-BOLD and 21 for BOLD-1b, respectively, to ensure that their respective total number of treated patients, which includes the backfilled patients in BF-BOLD and the additional 10 patients in BOLD-1b, are closely aligned. These tables indicate that the overall patient counts for both methods are similar, and it is evident that both the MTD and OBD accuracy levels for BF-BOLD exceed those of BOLD-1b, irrespective of the gap. This is especially true if we consider if MTD ≥ 1, which is interesting when it is believed that at least one of the doses will have acceptable toxicity. Interestingly, for the bottom state, BOLD-1b performs better. This could be due to the expansion phase where concentrated administration of the lowest dose can help resolve an initial misclassification of MTD as the lowest dose when it is actually too toxic. BOLD has relative difficulty in identifying when all doses are too toxic and the bottom state is true. Hence, this can be a factor to keep in mind in favor of BOLD-1b if there is a concern that the bottom state is viable. Also, the overdose rate for BF-BOLD is lower than that of BOLD-1b. These findings highlight the overall advantages of using backfilling in BF-BOLD to assess safety and efficacy with an overlapping activity marker in comparison to using BOLD with an added dose-expansion trial (i.e., BOLD-1b).

**Table 3:** Comparative analysis of BF-BOLD vs. BOLD-1b on accuracy (MTD and OBD) and efficiency (patient count) with a 5-dose model, where an activity plateau is present. A random number of backfilled patients (1 to 3) in BF-BOLD receive the anti-cover dose corresponding to the current dose selected for dose finding per stage, and 10 additional patients are treated with the identified MTD in BOLD-1b with no backfilling implemented. *N*_*max*_ are set to be 24 for BF-BOLD and 21 for BOLD-1b, respectively, ensuring that their overall numbers of treated patients are closely aligned. “Avg.” denotes the average for all the 5 MTD levels and “(≥1)” denotes the average for the MTD levels that are ≥ 1. All the other design characteristics and parameter settings are identical to those in Table 1.

| Gap<br>(MTD<br>minus<br>OBD) | MTD<br>level | Accuracy of identification (%) |  |  |  |  |  |  |  | Number of treated patients |  |
| --- | --- | --- | --- | --- | --- | --- | --- | --- | --- | --- | --- |
|  |  | MTD |  | OBD |  |  |  |  |  |  |  |
|  |  |  |  | Overall |  | When identified MTD = true<br>MTD |  | When identified MTD > true<br>MTD |  |  |  |
|  |  | BF-BOLD | BOLD-1b | BF-BOLD | BOLD-1b | BF-BOLD | BOLD-1b | BF-BOLD | BOLD-1b | BF-BOLD | BOLD-1b |
| 0 | <1 | 54.80 (13.84) | 66.32 (15.07) | -- | -- | -- | -- | -- | -- | 14.41 (2.28) | 18.18 (3.02) |
|  | 1 | 67.56 (4.29) | 58.72 (5.50) | 78.32 (4.22) | 73.56 (4.41) | 94.80 (2.51) | 97.72 (1.89) | 66.45 (11.31) | 56.70 (7.88) | 21.86 (0.84) | 27.05 (0.67) |
|  | 2 | 59.36 (9.43) | 54.38 (7.44) | 43.86 (7.01) | 42.36 (6.14) | 62.37 (7.10) | 60.21 (6.94) | 41.90 (8.93) | 36.64 (9.15) | 27.96 (0.92) | 29.82 (0.47) |
|  | 3 | 49.80 (10.49) | 48.82 (7.85) | 37.76 (8.59) | 37.26 (8.01) | 59.08 (5.92) | 60.25 (8.58) | 41.28 (10.67) | 31.90 (11.92) | 29.91 (1.28) | 30.56 (0.29) |
|  | 4 | 45.72 (7.45) | 45.44 (6.42) | 32.78 (5.95) | 30.80 (5.64) | 56.85 (7.52) | 56.02 (8.33) | 34.38 (13.37) | 30.61 (12.32) | 30.46 (1.39) | 30.81 (0.14) |
|  | 5 | 56.78 (9.25) | 51.36 (8.74) | 33.40 (7.46) | 28.42 (6.46) | 59.11 (7.43) | 55.33 (8.06) | -- | -- | 29.81 (1.67) | 30.90 (0.12) |
|  | >5 | 85.48 (8.19) | 76.32 (10.12) | 36.88 (10.49) | 26.92 (8.82) | 46.59 (11.57) | 35.43 (11.01) | -- | -- | 29.74 (1.46) | 30.96 (0.05) |
|  | Avg. | 59.93 (8.99) | 57.34 (8.73) | 43.83 (7.29) | 39.89 (6.58) | 63.13 (7.01) | 60.83 (7.47) | 46.00 (11.07) | 38.96 (10.32) | 26.30 (1.41) | 28.33 (0.68) |
|  | (≥1) | 60.78 (8.18) | 55.84 (7.68) | 43.83 (7.29) | 39.89 (6.58) | 63.13 (7.01) | 60.83 (7.47) | 46.00 (11.07) | 38.96 (10.32) | 28.29 (1.26) | 30.02 (0.29) |
| 1 | <1 | 54.80 (13.84) | 66.32 (15.07) | -- | -- | -- | -- | -- | -- | 14.41 (2.28) | 18.18 (3.02) |
|  | 1 | 67.56 (4.29) | 58.72 (5.50) | -- | -- | -- | -- | -- | -- | 21.86 (0.84) | 27.05 (0.67) |
|  | 2 | 59.48 (9.52) | 54.38 (7.44) | 68.16 (5.80) | 63.54 (5.02) | 65.75 (7.09) | 60.35 (6.80) | 52.59 (11.14) | 49.16 (11.32) | 28.51 (0.78) | 29.82 (0.47) |
|  | 3 | 50.14 (10.03) | 48.80 (7.98) | 43.20 (5.62) | 37.20 (5.77) | 43.50 (5.79) | 34.33 (8.22) | 29.71 (10.32) | 28.02 (10.45) | 31.39 (1.19) | 30.55 (0.32) |
|  | 4 | 45.32 (7.70) | 44.70 (6.38) | 38.38 (5.38) | 35.10 (5.56) | 40.38 (7.81) | 33.28 (6.63) | 27.24 (11.26) | 23.21 (11.74) | 31.87 (0.97) | 30.82 (0.17) |
|  | 5 | 56.06 (10.39) | 48.56 (10.53) | 36.62 (6.58) | 34.98 (6.24) | 37.91 (6.22) | 33.03 (7.52) | -- | -- | 31.25 (1.37) | 30.90 (0.10) |
|  | >5 | 84.58 (8.79) | 77.74 (11.00) | 49.96 (8.35) | 42.40 (7.82) | 58.90 (6.42) | 54.61 (7.24) | -- | -- | 30.94 (1.12) | 30.97 (0.06) |
|  | Avg. | 59.71 (9.22) | 57.03 (9.13) | 47.26 (6.35) | 42.64 (6.08) | 49.29 (6.66) | 43.12 (7.28) | 36.51 (10.91) | 33.46 (11.17) | 27.17 (1.22) | 28.32 (0.69) |
|  | (≥1) | 60.52 (8.45) | 55.48 (8.14) | 47.26 (6.35) | 42.64 (6.08) | 49.29 (6.66) | 43.12 (7.28) | 36.51 (10.91) | 33.46 (11.17) | 29.30 (1.05) | 30.02 (0.30) |
| 2 | <1 | 54.80 (13.84) | 66.32 (15.07) | -- | -- | -- | -- | -- | -- | 14.41 (2.28) | 18.18 (3.02) |
|  | 1 | 67.56 (4.29) | 58.72 (5.50) | -- | -- | -- | -- | -- | -- | 21.86 (0.84) | 27.05 (0.67) |
|  | 2 | 59.48 (9.52) | 54.38 (7.44) | -- | -- | -- | -- | -- | -- | 28.51 (0.78) | 29.82 (0.47) |
|  | 3 | 49.12 (8.93) | 48.80 (7.98) | 57.54 (5.26) | 51.50 (6.00) | 52.25 (8.35) | 46.89 (6.73) | 51.44 (8.93) | 46.82 (10.13) | 31.82 (1.12) | 30.55 (0.32) |
|  | 4 | 44.54 (7.08) | 46.00 (6.54) | 35.10 (4.94) | 30.62 (4.94) | 28.96 (7.36) | 25.27 (6.84) | 28.77 (10.28) | 22.07 (10.95) | 33.20 (0.76) | 30.81 (0.14) |
|  | 5 | 55.30 (10.73) | 50.52 (9.86) | 33.92 (5.75) | 29.56 (4.65) | 28.93 (7.71) | 22.29 (5.97) | -- | -- | 32.63 (1.22) | 30.89 (0.10) |
|  | >5 | 84.02 (8.90) | 76.38 (9.87) | 37.72 (4.97) | 32.40 (4.73) | 37.53 (5.29) | 30.42 (5.56) | -- | -- | 32.48 (1.06) | 30.95 (0.07) |
|  | Avg. | 59.26 (9.04) | 57.30 (8.89) | 41.07 (5.23) | 36.02 (5.08) | 36.92 (7.18) | 31.22 (6.28) | 40.10 (9.60) | 34.45 (10.54) | 27.84 (1.15) | 28.32 (0.69) |
|  | (≥1) | 60.00 (8.24) | 55.80 (7.86) | 41.07 (5.23) | 36.02 (5.08) | 36.92 (7.18) | 31.22 (6.28) | 40.10 (9.60) | 34.45 (10.54) | 30.08 (0.96) | 30.01 (0.30) |

## 6. Discussion

The BOLD algorithm can facilitate the identification of an OBD through the implementation of the backfill method. The benefits of backfilling are clear, as it allows for identifying a dose that is below the MTD while still maintaining the drug’s effectiveness. Such a dose may be more favorable than the MTD, as it presents a lower risk of DLTs and may improve practical sustainability of the treatment. Our method for determining the OBD with binary outcomes is straightforward and does not require a complex statistical model.

However, we do note that BF-BOLD is often not accurate enough to be the sole basis for OBD identification. OBD identification is a challenging classification problem, as it involves identifying a plateau or changepoint in response activity probabilities, and an OBD’s activity rate is not clearly differentiated with those for doses above it. This is exacerbated by the small sample nature of dose finding trials. Backfilling with 1-3 patients per stage (or taking as many patients as available to backfill) definitely improves performance, especially in OBD classification performance but also incrementally in MTD identification accuracy. Further, as in Tables S2 and S3, the performance with OBD identification improves depending on the discrimination between activity rates for the OBD and the next lowest dose (i.e. increased *δ*^*a*^). To determine OBD in practice, it will be prudent to use in conjunction all available data, in addition to statistical results from BF-BOLD, such as from pharmacokinetic and pharmacodynamic modeling, or other measurable factors that can reflect treatment engagement.

Note in Table S6, specificity is high in terms of detecting that activity rates for dose levels at or below MTD are less than a given threshold. This is important, as lack of observed activity in a dose-finding backfill design may be able to provide useful information about the futility of efficacy for doses considered at or below the target toxicity threshold without the need for a dose expansion study.

Another appealing feature of backfill designs, as originally purposed, is that patients assigned backfill doses should be receiving doses established as relative safe already. In terms of overdose rates, use of BF-BOLD leads to lower overdose rates than BOLD itself. Moreover, BOLD comes with an overdose control feature, in terms of reducing the *τ* parameter value below 0.50, as in (3). BF-BOLD also can adopt more stringent overdose control, as illustrated in Tables S4 and S5. This combination leads to a powerful control of overdosing.

In Figures 3 and S2, and Tables S7 and S8, performance comparisons are given between BF-BOLD and BF-BOIN. These are respective backfill versions of BOLD and BOIN, which was extensively studied in Wang and Tatsuoka (2026). Similarly as in prior study, we see for the most part, BF-BOLD has just as good or better performance in MTD accuracy, with a slightly smaller number of patients required on average. For the bottom state, BF-BOIN does perform better, but performs worse for accurately identifying the MTD when it is at higher dose levels. As Figure 3 shows, BF-BOLD also performs better as the discrimination between the DLT rates (i.e. upper *δ* values) of the MTD and its cover dose increases. OBD classification was not reported in the BF-BOIN app. We do note that higher MTD accuracy is important for OBD accuracy, since the possible dose levels under consideration as OBD depends on the MTD identification. In Wang and Tatsuoka (2026), BOLD had a relatively higher tendency for overestimation of MTD. The chance for recovery of OBD is less than if the identified MTD is correct, but it is still high, depending on the gap size. BOIN was found to have a relatively higher tendency to underestimate the MTD. If gap = 0 between MTD and OBD, this means that the OBD identification is necessarily incorrect. Hence, in OBD identification, underestimation of the MTD can be problematic. BOLD-based backfilling with the goal of identifying an OBD may be preferable for these reasons, especially if it is believed that at least one of the doses should be safe and the bottom state is unlikely (MTD ≥ 1).

Finally, in settings such as neoadjuvant studies where clinically valid activation markers can be obtained that will also serve as efficacy outcomes in a dose expansion trial, BF-BOLD can outperform BOLD plus a dose expansion phase (BOLD-1b) where the identified MTD is the dose being administered. See Tables 3 and S9. A potential issue with such a dose expansion design is that the MTD is often incorrectly identified in the dose-finding phase. A Phase 1b trial can allow for further testing of safety while collecting initial information on efficacy. However, fixing a single dose too early can lead to somewhat higher overdosing if the MTD is overestimated and collecting data from the same dose may limit correction of misidentification. Backfilling is more flexible and remains adaptive in dose selection, to enhance estimation of DLT rates across doses and discrimination between dose levels. An exception is when the bottom state is true and the lowest dose is incorrectly identified as MTD. Focused administration of the lowest dose, as in a single dose Phase 1b study, can help correct the misidentification. However, generally BF-BOLD outperforms BOLD-1b in other scenarios.

## 7. Conclusion

The BF-BOLD design framework is primarily aimed at helping to determine whether an optimal biological dose (OBD) exists that is lower than MTD. It involves collecting activation response data as well as toxicity data. Backfill designs deserve serious consideration in settings such as neoadjuvant studies when activation marker responses can be equivalent to clinical outcomes such as pathological response based on tissue analysis. Backfill designs also reduce overdose rates, so they can be viewed as overdose control by themselves, or even more effectively in conjunction with BOLD’s overdose control approach. These designs also have high specificity in identifying when activity rates are below minimum thresholds, which can be useful information concerning the viability of an efficacy study with the doses deemed safe. Moreover, the proposed Bayesian approach is computationally simple, avoiding the need for simulation. This adaptability and ease of use underscores the extensive capabilities of BF-BOLD designs for Phase I clinical trials.

## Data Availability

All data produced in the present study are available upon reasonable request to the authors

## Acknowledgements

This work was supported in part by National Science Foundation grant 2545715 and National Cancer Institute grants P30 CA134274-10 (Greenebaum Comprehensive Cancer Center) and P30 CA043703-34 (Case Comprehensive Cancer Center).

