## Supplemental Materials for "Backfill Bayesian Ordered Lattice Design for Phase I Clinical Trials"

### Supplementary Material

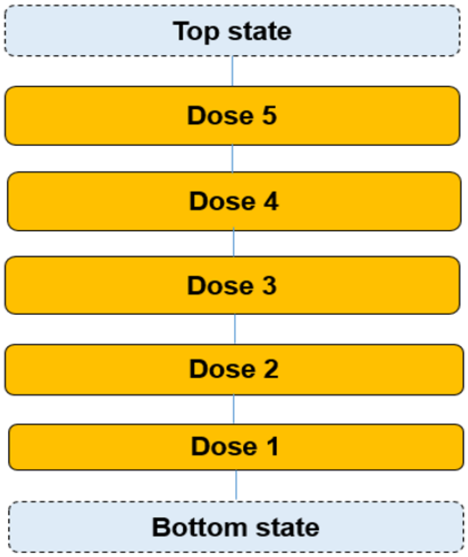


Figure S1: Hasse diagram of 5-dose lattice with linear order, where the bottom state represents that a MTD is below the minimum dose, and the top state signifies that the MTD exceeds the maximum dose. Classification is to one state, and classification to a specific dose level indicates that the corresponding dose is identified as the MTD.

| Gap (MTD minus OBD) | MTD level | Accuracy of identification (SD) | | | | Number of treated patients (SD) | | Overdose rate | | Underestimation rate | | Overestimation rate | |
|---|---|---|---|---|---|---|---|---|---|---|---|---|---|
| | | MTD | OBD | | | | | | | | | | |
| | | | Overall | When identified MTD = true MTD | When identified MTD > true MTD | Overall | Backfill only | Overall | Backfill only | MTD | OBD | MTD | OBD |
| 0 | <1 | 55.00 (13.99) | -- | -- | -- | 14.48 (2.38) | 1.55 (0.68) | 0.19 | 0.17 | -- | -- | 0.45 | -- |
| | 1 | 69.06 (4.67) | 79.00 (3.98) | 95.66 (2.14) | 64.34 (10.95) | 22.45 (0.93) | 4.54 (0.45) | 0.38 | 0.23 | 0.11 | 0.14 | 0.20 | 0.07 |
| | 2 | 60.52 (8.95) | 45.26 (7.30) | 62.04 (6.04) | 41.27 (11.20) | 29.49 (0.96) | 8.67 (0.68) | 0.25 | 0.10 | 0.21 | 0.50 | 0.19 | 0.05 |
| | 3 | 52.64 (10.86) | 39.20 (7.91) | 60.52 (6.30) | 38.54 (10.83) | 33.13 (1.41) | 9.45 (1.13) | 0.20 | 0.07 | 0.28 | 0.55 | 0.19 | 0.06 |
| | 4 | 48.28 (8.51) | 35.02 (6.91) | 58.22 (7.86) | 40.25 (12.51) | 35.24 (1.42) | 9.46 (1.36) | 0.13 | 0.00 | 0.35 | 0.60 | 0.17 | 0.05 |
| | 5 | 59.56 (9.35) | 36.26 (7.36) | 60.75 (6.65) | -- | 34.86 (1.73) | 8.35 (1.76) | -- | -- | 0.40 | 0.64 | -- | -- |
| | >5 | 89.04 (7.00) | 42.48 (10.20) | 47.78 (11.42) | -- | 33.52 (1.72) | 7.36 (1.81) | -- | -- | 0.11 | 0.58 | -- | -- |
| | **Avg.** | **62.01 (9.05)** | **46.20 (7.28)** | **64.16 (6.74)** | **46.10 (11.37)** | **29.02 (1.51)** | **7.05 (1.12)** | **0.23** | **0.11** | **0.24** | **0.50** | **0.24** | **0.06** |
| 1 | <1 | 55.00 (13.99) | -- | -- | -- | 14.48 (2.38) | 1.55 (0.68) | 0.19 | 0.17 | -- | -- | 0.45 | -- |
| | 1 | 69.06 (4.67) | -- | -- | -- | 22.45 (0.93) | 4.54 (0.45) | 0.38 | 0.23 | 0.11 | -- | 0.20 | -- |
| | 2 | 61.20 (9.38) | 71.10 (5.00) | 69.67 (6.96) | 52.95 (10.61) | 30.06 (0.95) | 9.30 (0.72) | 0.25 | 0.09 | 0.20 | 0.03 | 0.19 | 0.26 |
| | 3 | 52.84 (9.99) | 43.24 (5.18) | 42.89 (5.71) | 30.88 (10.95) | 34.79 (1.18) | 11.24 (0.86) | 0.19 | 0.06 | 0.28 | 0.34 | 0.19 | 0.23 |
| | 4 | 47.74 (8.24) | 40.86 (5.19) | 43.62 (8.07) | 27.33 (9.08) | 37.00 (0.97) | 11.23 (0.79) | 0.13 | 0.00 | 0.34 | 0.38 | 0.18 | 0.21 |
| | 5 | 59.50 (11.44) | 39.14 (5.90) | 39.81 (5.25) | -- | 36.76 (1.47) | 10.40 (1.25) | -- | -- | 0.41 | 0.42 | -- | 0.19 |
| | >5 | 89.78 (7.07) | 54.84 (7.09) | 61.03 (5.54) | -- | 35.63 (1.07) | 9.42 (1.12) | -- | -- | 0.10 | 0.45 | -- | -- |
| | **Avg.** | **62.16 (9.25)** | **49.84 (5.67)** | **51.40 (6.31)** | **37.05 (10.21)** | **30.17 (1.28)** | **8.24 (0.84)** | **0.23** | **0.11** | **0.24** | **0.33** | **0.24** | **0.22** |
| 2 | <1 | 55.00 (13.99) | -- | -- | -- | 14.48 (2.38) | 1.55 (0.68) | 0.19 | 0.17 | -- | -- | 0.45 | -- |
| | 1 | 69.06 (4.67) | -- | -- | -- | 22.45 (0.93) | 4.54 (0.45) | 0.38 | 0.23 | 0.11 | -- | 0.20 | -- |
| | 2 | 61.20 (9.38) | -- | -- | -- | 30.06 (0.95) | 9.30 (0.72) | 0.25 | 0.09 | 0.20 | -- | 0.19 | -- |
| | 3 | 52.18 (10.39) | 57.26 (6.06) | 52.61 (7.50) | 51.09 (10.70) | 35.14 (1.28) | 11.63 (0.88) | 0.19 | 0.05 | 0.29 | 0.01 | 0.19 | 0.42 |
| | 4 | 48.48 (8.56) | 37.16 (4.93) | 33.42 (6.73) | 28.34 (10.09) | 38.48 (1.07) | 12.77 (0.68) | 0.12 | 0.00 | 0.34 | 0.29 | 0.17 | 0.34 |
| | 5 | 58.50 (10.01) | 34.56 (4.73) | 29.09 (6.26) | -- | 38.27 (1.19) | 11.95 (0.92) | -- | -- | 0.42 | 0.32 | -- | 0.33 |
| | >5 | 88.92 (7.27) | 40.26 (5.27) | 40.92 (5.71) | -- | 37.55 (0.93) | 11.40 (0.92) | -- | -- | 0.11 | 0.34 | -- | 0.26 |
| | **Avg.** | **61.91 (9.18)** | **42.31 (5.25)** | **39.01 (6.55)** | **39.71 (10.39)** | **30.92 (1.25)** | **9.02 (0.75)** | **0.22** | **0.11** | **0.24** | **0.24** | **0.24** | **0.34** |

Table S1: Summary of identification accuracy, number of treated patients, overdose rate, underestimation rate, and overestimation rate of BF-BOLD design with 5-dose lattice at $\phi = 0.25$, $\phi^a = 0.5$, true activity rate of OBD $= 0.7$, upper $\delta$ (for DLT rates) $= 0.15$, lower $\delta^a$ (for activity rates) $= 0.15$, and $N_{max} = 30$, utilizing random scenarios of true DLT rates and activity rates, while OBD being zero, one or two levels below the true MTD. An activity plateau is present, and a random number of backfilled patients (up to 3) receive the anti-cover dose corresponding to the current dose in dose-finding. A 10% activity rate trade-off is utilized for the identification of OBD.

| Gap (MTD minus OBD) | MTD level | Accuracy of identification (SD) | | | | Number of treated patients (SD) | | Overdose rate | | Underestimation rate | | Overestimation rate | |
|---|---|---|---|---|---|---|---|---|---|---|---|---|---|
| | | MTD | OBD | | | | | | | | | | |
| | | | Overall | When identified MTD = true MTD | When identified MTD > true MTD | Overall | Backfill only | Overall | Backfill only | MTD | OBD | MTD | OBD |
| 0 | <1 | 54.82 (14.00) | -- | -- | -- | 14.20 (2.16) | 1.43 (0.60) | 0.19 | 0.15 | -- | -- | 0.45 | -- |
| | 1 | 66.02 (4.75) | 77.56 (4.39) | 94.79 (2.44) | 66.61 (11.06) | 21.06 (0.73) | 3.93 (0.36) | 0.37 | 0.21 | 0.12 | 0.15 | 0.22 | 0.07 |
| | 2 | 56.14 (8.61) | 49.50 (7.68) | 69.81 (6.52) | 45.35 (11.00) | 25.65 (0.85) | 6.59 (0.74) | 0.23 | 0.08 | 0.21 | 0.43 | 0.22 | 0.07 |
| | 3 | 47.96 (9.06) | 40.96 (7.95) | 64.91 (6.94) | 40.34 (9.37) | 26.59 (0.98) | 6.38 (0.81) | 0.16 | 0.04 | 0.28 | 0.51 | 0.24 | 0.08 |
| | 4 | 42.80 (6.50) | 33.54 (6.53) | 59.15 (6.81) | 41.49 (10.59) | 26.20 (1.08) | 5.48 (1.03) | 0.09 | 0.00 | 0.37 | 0.60 | 0.20 | 0.06 |
| | 5 | 50.64 (11.01) | 29.94 (8.00) | 58.88 (7.39) | -- | 25.33 (1.28) | 4.47 (1.22) | -- | -- | 0.49 | 0.70 | -- | -- |
| | >5 | 79.44 (9.68) | 29.02 (7.13) | 36.63 (8.35) | -- | 25.15 (1.08) | 4.20 (1.07) | -- | -- | 0.21 | 0.71 | -- | -- |
| | **Avg.** | **56.83 (9.09)** | **43.42 (6.95)** | **64.03 (6.41)** | **48.45 (10.50)** | **23.45 (1.17)** | **4.64 (0.83)** | **0.21** | **0.10** | **0.28** | **0.52** | **0.27** | **0.07** |
| 1 | <1 | 54.82 (14.00) | -- | -- | -- | 14.20 (2.16) | 1.43 (0.60) | 0.19 | 0.15 | -- | -- | 0.45 | -- |
| | 1 | 66.02 (4.75) | -- | -- | -- | 21.06 (0.73) | 3.93 (0.36) | 0.37 | 0.21 | 0.12 | -- | 0.22 | -- |
| | 2 | 57.50 (9.18) | 69.44 (5.18) | 67.24 (6.80) | 55.36 (9.99) | 26.65 (0.80) | 7.58 (0.75) | 0.23 | 0.06 | 0.20 | 0.03 | 0.22 | 0.27 |
| | 3 | 48.26 (9.86) | 46.42 (6.23) | 46.75 (5.55) | 33.70 (11.02) | 28.28 (1.04) | 8.11 (0.82) | 0.15 | 0.03 | 0.28 | 0.29 | 0.24 | 0.24 |
| | 4 | 42.58 (8.21) | 43.68 (5.29) | 46.54 (7.33) | 33.57 (9.40) | 27.73 (0.75) | 7.03 (0.68) | 0.08 | 0.00 | 0.37 | 0.35 | 0.20 | 0.22 |
| | 5 | 50.54 (10.53) | 40.92 (7.75) | 43.87 (7.48) | -- | 26.58 (1.12) | 5.75 (1.02) | -- | -- | 0.49 | 0.43 | -- | 0.16 |
| | >5 | 78.74 (10.18) | 48.40 (7.33) | 61.52 (5.22) | -- | 26.10 (0.94) | 5.15 (0.92) | -- | -- | 0.21 | 0.52 | -- | -- |
| | **Avg.** | **56.92 (9.53)** | **49.77 (6.35)** | **53.18 (6.48)** | **40.88 (10.14)** | **24.37 (1.08)** | **5.57 (0.74)** | **0.20** | **0.09** | **0.28** | **0.32** | **0.27** | **0.22** |
| 2 | <1 | 54.82 (14.00) | -- | -- | -- | 14.20 (2.16) | 1.43 (0.60) | 0.19 | 0.15 | -- | -- | 0.45 | -- |
| | 1 | 66.02 (4.75) | -- | -- | -- | 21.06 (0.73) | 3.93 (0.36) | 0.37 | 0.21 | 0.12 | -- | 0.22 | -- |
| | 2 | 57.50 (9.18) | -- | -- | -- | 26.65 (0.80) | 7.58 (0.75) | 0.23 | 0.06 | 0.20 | -- | 0.22 | -- |
| | 3 | 46.62 (8.67) | 56.94 (5.89) | 52.49 (8.47) | 49.82 (11.06) | 28.64 (1.03) | 8.53 (0.71) | 0.15 | 0.03 | 0.30 | 0.01 | 0.24 | 0.42 |
| | 4 | 43.44 (7.37) | 39.36 (5.55) | 34.79 (7.77) | 30.37 (12.55) | 29.13 (0.63) | 8.49 (0.49) | 0.08 | 0.00 | 0.37 | 0.26 | 0.20 | 0.34 |
| | 5 | 52.14 (11.62) | 40.30 (5.51) | 32.85 (7.74) | -- | 28.10 (0.86) | 7.28 (0.73) | -- | -- | 0.48 | 0.29 | -- | 0.31 |
| | >5 | 77.86 (10.74) | 43.00 (5.16) | 42.34 (5.83) | -- | 27.39 (0.83) | 6.46 (0.79) | -- | -- | 0.22 | 0.32 | -- | 0.25 |
| | **Avg.** | **56.91 (9.47)** | **44.90 (5.53)** | **40.62 (7.45)** | **40.10 (11.81)** | **25.02 (1.00)** | **6.24 (0.63)** | **0.20** | **0.09** | **0.28** | **0.22** | **0.27** | **0.33** |

Table S2: Summary of identification accuracy, number of treated patients, overdose rate, underestimation rate, and overestimation rate of BF-BOLD design with 5-dose lattice, $N_{max} = 21$, and lower $\delta^a$ for random scenarios of true activity rates set to be 0.2. All the other design characteristics, parameter settings, and analysis procedures are identical to those in Table S1.

| Gap (MTD minus OBD) | MTD level | Accuracy of identification (SD) | | | | Number of treated patients (SD) | | Overdose rate | | Underestimation rate | | Overestimation rate | |
|---|---|---|---|---|---|---|---|---|---|---|---|---|---|
| | | MTD | OBD | | | Overall | Backfill only | Overall | Backfill only | MTD | OBD | MTD | OBD |
| | | | Overall | When identified MTD = true MTD | When identified MTD > true MTD | | | | | | | | |
| 0 | <1 | 55.00 (13.99) | -- | -- | -- | 14.48 (2.38) | 1.55 (0.68) | 0.19 | 0.17 | -- | -- | 0.45 | -- |
| | 1 | 69.06 (4.67) | 79.00 (3.98) | 95.66 (2.14) | 64.34 (10.95) | 22.45 (0.93) | 4.54 (0.45) | 0.38 | 0.23 | 0.11 | 0.14 | 0.20 | 0.07 |
| | 2 | 62.38 (8.92) | 53.66 (6.91) | 72.04 (5.27) | 48.99 (9.56) | 29.17 (0.93) | 8.35 (0.73) | 0.25 | 0.10 | 0.20 | 0.41 | 0.18 | 0.06 |
| | 3 | 53.82 (9.40) | 46.70 (9.07) | 71.14 (6.26) | 45.67 (11.14) | 32.59 (1.17) | 8.96 (1.00) | 0.20 | 0.07 | 0.28 | 0.48 | 0.18 | 0.05 |
| | 4 | 48.90 (7.63) | 40.78 (7.34) | 67.06 (7.74) | 48.34 (15.99) | 34.50 (1.27) | 8.79 (1.19) | 0.13 | 0.00 | 0.35 | 0.54 | 0.16 | 0.05 |
| | 5 | 59.58 (10.13) | 42.02 (7.96) | 70.49 (5.58) | -- | 34.01 (1.56) | 7.49 (1.53) | -- | -- | 0.40 | 0.58 | -- | -- |
| | >5 | 89.86 (6.53) | 38.50 (8.83) | 42.83 (9.50) | -- | 32.60 (1.51) | 6.38 (1.53) | -- | -- | 0.10 | 0.62 | -- | -- |
| | **Avg.** | **62.66 (8.75)** | **50.11 (7.35)** | **69.87 (6.08)** | **51.83 (11.91)** | **28.54 (1.39)** | **6.58 (1.02)** | **0.23** | **0.11** | **0.24** | **0.46** | **0.23** | **0.06** |
| 1 | <1 | 55.00 (13.99) | -- | -- | -- | 14.48 (2.38) | 1.55 (0.68) | 0.19 | 0.17 | -- | -- | 0.45 | -- |
| | 1 | 69.06 (4.67) | -- | -- | -- | 22.45 (0.93) | 4.54 (0.45) | 0.38 | 0.23 | 0.11 | -- | 0.20 | -- |
| | 2 | 61.20 (9.38) | 71.10 (5.00) | 69.67 (6.96) | 52.95 (10.61) | 30.06 (0.95) | 9.30 (0.72) | 0.25 | 0.09 | 0.20 | 0.03 | 0.19 | 0.26 |
| | 3 | 52.88 (11.00) | 49.88 (5.05) | 49.93 (6.34) | 37.32 (11.72) | 34.52 (1.26) | 11.10 (0.93) | 0.19 | 0.06 | 0.29 | 0.26 | 0.19 | 0.24 |
| | 4 | 48.06 (8.44) | 45.86 (6.78) | 47.64 (8.64) | 34.11 (14.12) | 36.57 (0.97) | 10.82 (0.87) | 0.12 | 0.00 | 0.35 | 0.32 | 0.17 | 0.22 |
| | 5 | 59.24 (10.13) | 45.54 (6.15) | 46.42 (7.13) | -- | 36.40 (1.34) | 9.94 (1.22) | -- | -- | 0.41 | 0.36 | -- | 0.19 |
| | >5 | 90.16 (6.73) | 63.08 (7.02) | 69.90 (5.02) | -- | 34.77 (1.00) | 8.54 (1.02) | -- | -- | 0.10 | 0.37 | -- | -- |
| | **Avg.** | **62.23 (9.19)** | **55.09 (6.00)** | **56.71 (6.82)** | **41.46 (12.15)** | **29.89 (1.26)** | **7.97 (0.84)** | **0.22** | **0.11** | **0.24** | **0.27** | **0.24** | **0.23** |
| 2 | <1 | 55.00 (13.99) | -- | -- | -- | 14.48 (2.38) | 1.55 (0.68) | 0.19 | 0.17 | -- | -- | 0.45 | -- |
| | 1 | 69.06 (4.67) | -- | -- | -- | 22.45 (0.93) | 4.54 (0.45) | 0.38 | 0.23 | 0.11 | -- | 0.20 | -- |
| | 2 | 61.20 (9.38) | -- | -- | -- | 30.06 (0.95) | 9.30 (0.72) | 0.25 | 0.09 | 0.20 | -- | 0.19 | -- |
| | 3 | 52.18 (10.39) | 57.26 (6.06) | 52.61 (7.50) | 51.09 (10.70) | 35.14 (1.28) | 11.63 (0.88) | 0.19 | 0.05 | 0.29 | 0.01 | 0.19 | 0.42 |
| | 4 | 48.54 (8.16) | 39.86 (5.51) | 34.88 (5.98) | 33.14 (11.93) | 38.28 (0.88) | 12.60 (0.61) | 0.12 | 0.00 | 0.35 | 0.25 | 0.17 | 0.35 |
| | 5 | 58.90 (9.97) | 38.30 (3.60) | 31.11 (6.28) | -- | 37.95 (1.06) | 11.66 (0.88) | -- | -- | 0.41 | 0.26 | -- | 0.36 |
| | >5 | 89.02 (7.10) | 45.40 (6.12) | 45.58 (6.19) | -- | 37.22 (0.74) | 10.99 (0.82) | -- | -- | 0.11 | 0.26 | -- | 0.29 |
| | **Avg.** | **61.99 (9.09)** | **45.21 (5.32)** | **41.05 (6.49)** | **42.12 (11.32)** | **30.80 (1.17)** | **8.89 (0.72)** | **0.22** | **0.11** | **0.24** | **0.19** | **0.24** | **0.36** |

Table S3: Summary of identification accuracy, number of treated patients, overdose rate, underestimation rate, and overestimation rate of BF-BOLD design with 5-dose lattice, $N_{max} = 30$, and lower $\delta^a$ for random scenarios of true activity rates set to be 0.2. All the other design characteristics, parameter settings, and analysis procedures are identical to those in Table S1.

| Gap (MTD minus OBD) | MTD level | Accuracy of identification (SD) | | | | Number of treated patients (SD) | | Overdose rate | | Underestimation rate | | Overestimation rate | |
|---|---|---|---|---|---|---|---|---|---|---|---|---|---|
| | | MTD | OBD | | | Overall | Backfill only | Overall | Backfill only | MTD | OBD | MTD | OBD |
| | | | Overall | When identified MTD = true MTD | When identified MTD > true MTD | | | | | | | | |
| 0 | <1 | 54.70 (13.25) | -- | -- | -- | 13.74 (1.87) | 1.15 (0.51) | 0.15 | 0.13 | -- | -- | 0.45 | -- |
| | 1 | 68.94 (4.84) | 78.96 (4.08) | 95.26 (2.43) | 66.89 (11.73) | 20.11 (0.51) | 3.29 (0.28) | 0.31 | 0.19 | 0.11 | 0.15 | 0.20 | 0.06 |
| | 2 | 58.16 (10.04) | 43.08 (8.29) | 61.08 (6.05) | 39.41 (11.77) | 25.19 (0.95) | 6.42 (0.81) | 0.19 | 0.06 | 0.23 | 0.51 | 0.19 | 0.06 |
| | 3 | 44.60 (9.34) | 34.06 (8.65) | 58.45 (7.95) | 38.09 (11.85) | 26.25 (1.13) | 6.36 (0.93) | 0.13 | 0.03 | 0.35 | 0.60 | 0.20 | 0.06 |
| | 4 | 37.02 (8.27) | 26.40 (5.50) | 52.90 (7.51) | 38.25 (13.32) | 26.13 (1.28) | 5.69 (1.17) | 0.08 | 0.00 | 0.45 | 0.68 | 0.18 | 0.05 |
| | 5 | 49.28 (10.69) | 26.78 (6.72) | 54.50 (8.26) | -- | 25.46 (1.49) | 4.74 (1.39) | -- | -- | 0.51 | 0.73 | -- | -- |
| | >5 | 75.56 (11.81) | 32.12 (8.77) | 42.48 (9.50) | -- | 25.46 (1.25) | 4.58 (1.22) | -- | -- | 0.24 | 0.68 | -- | -- |
| | **Avg.** | **55.47 (9.75)** | **40.23 (7.00)** | **60.78 (6.95)** | **45.66 (12.17)** | **23.19 (1.21)** | **4.60 (0.90)** | **0.17** | **0.08** | **0.32** | **0.56** | **0.24** | **0.06** |
| 1 | <1 | 54.70 (13.25) | -- | -- | -- | 13.74 (1.87) | 1.15 (0.51) | 0.15 | 0.13 | -- | -- | 0.45 | -- |
| | 1 | 68.94 (4.84) | -- | -- | -- | 20.11 (0.51) | 3.29 (0.28) | 0.31 | 0.19 | 0.11 | -- | 0.20 | -- |
| | 2 | 58.34 (10.57) | 71.18 (4.73) | 68.72 (5.05) | 52.89 (11.34) | 25.67 (1.04) | 6.99 (0.88) | 0.19 | 0.06 | 0.24 | 0.03 | 0.18 | 0.26 |
| | 3 | 44.60 (9.94) | 42.66 (6.71) | 43.07 (7.78) | 29.35 (9.78) | 27.69 (1.12) | 7.87 (0.84) | 0.13 | 0.03 | 0.36 | 0.37 | 0.19 | 0.20 |
| | 4 | 37.48 (8.61) | 37.32 (5.72) | 42.14 (8.44) | 30.69 (9.05) | 27.38 (1.03) | 6.99 (0.89) | 0.07 | 0.00 | 0.45 | 0.45 | 0.18 | 0.17 |
| | 5 | 47.68 (10.97) | 31.78 (6.70) | 35.70 (7.46) | -- | 26.72 (1.34) | 6.06 (1.21) | -- | -- | 0.52 | 0.53 | -- | 0.15 |
| | >5 | 75.52 (11.01) | 42.50 (8.19) | 56.23 (6.62) | -- | 26.40 (0.99) | 5.54 (0.96) | -- | -- | 0.24 | 0.58 | -- | -- |
| | **Avg.** | **55.32 (9.88)** | **45.09 (6.41)** | **49.17 (7.07)** | **37.64 (10.06)** | **23.96 (1.13)** | **5.41 (0.80)** | **0.17** | **0.08** | **0.32** | **0.39** | **0.24** | **0.20** |
| 2 | <1 | 54.70 (13.25) | -- | -- | -- | 13.74 (1.87) | 1.15 (0.51) | 0.15 | 0.13 | -- | -- | 0.45 | -- |
| | 1 | 68.94 (4.84) | -- | -- | -- | 20.11 (0.51) | 3.29 (0.28) | 0.31 | 0.19 | 0.11 | -- | 0.20 | -- |
| | 2 | 58.34 (10.57) | -- | -- | -- | 25.67 (1.04) | 6.99 (0.88) | 0.19 | 0.06 | 0.24 | -- | 0.18 | -- |
| | 3 | 45.36 (10.13) | 59.40 (7.55) | 53.49 (9.27) | 49.38 (10.22) | 28.05 (1.24) | 8.22 (0.88) | 0.13 | 0.03 | 0.35 | 0.02 | 0.20 | 0.39 |
| | 4 | 37.96 (7.79) | 37.80 (6.07) | 31.01 (8.36) | 28.25 (10.08) | 28.79 (0.80) | 8.42 (0.57) | 0.06 | 0.00 | 0.46 | 0.32 | 0.16 | 0.30 |
| | 5 | 47.26 (11.44) | 32.82 (5.82) | 26.81 (7.14) | -- | 27.96 (1.04) | 7.32 (0.87) | -- | -- | 0.53 | 0.39 | -- | 0.29 |
| | >5 | 74.66 (12.17) | 35.46 (5.66) | 36.70 (5.55) | -- | 27.56 (0.96) | 6.71 (0.89) | -- | -- | 0.25 | 0.42 | -- | 0.23 |
| | **Avg.** | **55.32 (10.03)** | **41.37 (6.28)** | **37.00 (7.58)** | **38.81 (10.15)** | **24.55 (1.06)** | **6.01 (0.70)** | **0.17** | **0.08** | **0.32** | **0.28** | **0.24** | **0.30** |

Table S4: Summary of identification accuracy, number of treated patients, overdose rate, underestimation rate, and overestimation rate of BF-BOLD design with 5-dose lattice, $N_{max} = 21$, and PPAT threshold parameter $\tau = 0.45$. All the other design characteristics, parameter settings, and analysis procedures are identical to those in Table S1.

| Gap (MTD minus OBD) | MTD level | Accuracy of identification (SD) | | | | Number of treated patients (SD) | | Overdose rate | | Underestimation rate | | Overestimation rate | |
|---|---|---|---|---|---|---|---|---|---|---|---|---|---|
| | | MTD | OBD | | | | | | | | | | |
| | | | Overall | When identified MTD = true MTD | When identified MTD > true MTD | Overall | Backfill only | Overall | Backfill only | MTD | OBD | MTD | OBD |
| 0 | <1 | 54.78 (13.38) | -- | -- | -- | 14.00 (2.03) | 1.23 (0.56) | 0.16 | 0.15 | -- | -- | 0.45 | -- |
| | 1 | 71.98 (4.41) | 80.14 (4.01) | 95.40 (2.91) | 67.52 (11.66) | 21.29 (0.92) | 3.80 (0.41) | 0.33 | 0.21 | 0.11 | 0.15 | 0.17 | 0.05 |
| | 2 | 60.88 (10.91) | 43.70 (8.52) | 61.34 (7.68) | 43.33 (13.46) | 27.75 (1.22) | 7.64 (0.89) | 0.20 | 0.08 | 0.25 | 0.52 | 0.15 | 0.04 |
| | 3 | 49.58 (11.64) | 35.26 (8.97) | 59.63 (7.45) | 40.72 (15.55) | 31.16 (1.36) | 8.58 (1.12) | 0.16 | 0.06 | 0.36 | 0.61 | 0.14 | 0.04 |
| | 4 | 42.08 (9.28) | 29.70 (7.00) | 58.79 (9.89) | 39.86 (15.84) | 33.08 (1.59) | 8.53 (1.43) | 0.10 | 0.00 | 0.45 | 0.66 | 0.12 | 0.04 |
| | 5 | 49.24 (9.66) | 29.92 (7.12) | 60.66 (7.39) | -- | 33.32 (1.97) | 7.71 (1.78) | -- | -- | 0.51 | 0.70 | -- | -- |
| | >5 | 81.88 (9.30) | 40.00 (10.00) | 48.92 (11.33) | -- | 32.88 (1.64) | 7.10 (1.69) | -- | -- | 0.18 | 0.60 | -- | -- |
| | **Avg.** | **58.63 (9.80)** | **43.12 (7.60)** | **64.13 (7.77)** | **47.86 (14.13)** | **27.64 (1.53)** | **6.37 (1.13)** | **0.19** | **0.10** | **0.31** | **0.54** | **0.21** | **0.04** |
| 1 | <1 | 54.78 (13.38) | -- | -- | -- | 14.00 (2.03) | 1.23 (0.56) | 0.16 | 0.15 | -- | -- | 0.45 | -- |
| | 1 | 71.98 (4.41) | -- | -- | -- | 21.29 (0.92) | 3.80 (0.41) | 0.33 | 0.21 | 0.11 | -- | 0.17 | -- |
| | 2 | 61.32 (11.28) | 71.68 (6.49) | 67.62 (6.57) | 50.66 (13.90) | 28.36 (1.11) | 8.29 (0.91) | 0.20 | 0.07 | 0.25 | 0.03 | 0.14 | 0.26 |
| | 3 | 50.02 (11.31) | 44.62 (5.84) | 44.41 (6.74) | 30.96 (13.98) | 32.83 (1.42) | 10.41 (1.06) | 0.15 | 0.05 | 0.36 | 0.36 | 0.14 | 0.19 |
| | 4 | 43.08 (9.23) | 40.12 (5.31) | 43.60 (8.08) | 31.18 (13.71) | 35.08 (1.42) | 10.45 (1.13) | 0.10 | 0.00 | 0.44 | 0.43 | 0.13 | 0.17 |
| | 5 | 49.90 (11.23) | 37.00 (8.39) | 42.39 (6.62) | -- | 35.17 (1.93) | 9.63 (1.46) | -- | -- | 0.50 | 0.48 | -- | 0.15 |
| | >5 | 82.02 (9.64) | 49.84 (7.71) | 60.75 (5.95) | -- | 34.87 (1.13) | 9.07 (1.17) | -- | -- | 0.18 | 0.50 | -- | -- |
| | **Avg.** | **59.01 (10.07)** | **48.65 (6.75)** | **51.75 (6.79)** | **37.60 (13.86)** | **28.80 (1.42)** | **7.55 (0.96)** | **0.19** | **0.10** | **0.31** | **0.36** | **0.21** | **0.19** |
| 2 | <1 | 54.78 (13.38) | -- | -- | -- | 14.00 (2.03) | 1.23 (0.56) | 0.16 | 0.15 | -- | -- | 0.45 | -- |
| | 1 | 71.98 (4.41) | -- | -- | -- | 21.29 (0.92) | 3.80 (0.41) | 0.33 | 0.21 | 0.11 | -- | 0.17 | -- |
| | 2 | 61.32 (11.28) | -- | -- | -- | 28.36 (1.11) | 8.29 (0.91) | 0.20 | 0.07 | 0.25 | -- | 0.14 | -- |
| | 3 | 50.52 (10.69) | 58.28 (6.68) | 51.51 (8.40) | 50.10 (12.23) | 33.37 (1.16) | 10.90 (0.90) | 0.15 | 0.04 | 0.35 | 0.01 | 0.15 | 0.41 |
| | 4 | 42.60 (9.84) | 39.00 (5.24) | 32.14 (6.34) | 30.83 (13.21) | 36.35 (1.32) | 11.99 (0.87) | 0.09 | 0.00 | 0.45 | 0.31 | 0.12 | 0.30 |
| | 5 | 49.36 (10.92) | 35.00 (4.49) | 29.17 (6.92) | -- | 36.81 (1.64) | 11.33 (1.22) | -- | -- | 0.51 | 0.35 | -- | 0.30 |
| | >5 | 81.52 (9.54) | 39.48 (5.53) | 41.34 (5.38) | -- | 36.61 (1.03) | 10.88 (1.08) | -- | -- | 0.18 | 0.36 | -- | 0.24 |
| | **Avg.** | **58.87 (10.01)** | **42.94 (5.48)** | **38.54 (6.76)** | **40.46 (12.72)** | **29.54 (1.32)** | **8.34 (0.85)** | **0.18** | **0.10** | **0.31** | **0.26** | **0.21** | **0.31** |

Table S5: Summary of identification accuracy, number of treated patients, overdose rate, underestimation rate, and overestimation rate of BF-BOLD design with 5-dose lattice, $N_{max} = 30$, and PPAT threshold parameter $\tau = 0.45$. All the other design characteristics, parameter settings, and analysis procedures are identical to those in Table S1.

| N_max for MTD trial | True activity rate of MTD | MTD level | Accuracy of identification (SD) | | | | Number of treated patients (SD) | | Overdose rate | | Under-estimate rate (MTD) | Overestimation rate | |
|---|---|---|---|---|---|---|---|---|---|---|---|---|---|
| | | | | No OBD identification | | | | | | | | | |
| | | | MTD | Overall | When identified MTD = true MTD | When identified MTD > true MTD | Overall | Backfill only | Overall | Backfill only | | MTD | OBD |
| 21 | 0.3 | <1 | 52.84 (14.19) | -- | -- | -- | 13.58 (1.73) | 0.46 (0.20) | 0.21 | 0.19 | -- | 0.47 | -- |
| | | 1 | 60.98 (4.65) | 95.50 (1.69) | 97.06 (1.88) | 90.75 (5.02) | 18.92 (0.45) | 1.13 (0.19) | 0.41 | 0.27 | 0.10 | 0.29 | 0.05 |
| | | 2 | 53.38 (6.51) | 93.00 (2.41) | 94.50 (3.23) | 85.59 (6.24) | 21.19 (0.44) | 1.48 (0.25) | 0.29 | 0.14 | 0.19 | 0.28 | 0.07 |
| | | 3 | 47.10 (8.75) | 92.06 (2.55) | 92.96 (4.01) | 82.28 (7.63) | 21.82 (0.46) | 1.35 (0.26) | 0.19 | 0.07 | 0.27 | 0.26 | 0.08 |
| | | 4 | 43.60 (6.61) | 90.54 (2.87) | 88.50 (5.36) | 77.78 (10.16) | 21.99 (0.34) | 1.21 (0.24) | 0.10 | 0.00 | 0.37 | 0.20 | 0.09 |
| | | 5 | 50.22 (9.03) | 89.78 (3.45) | 81.37 (6.15) | -- | 21.68 (0.27) | 0.80 (0.18) | -- | -- | 0.50 | -- | 0.10 |
| | | >5 | 78.72 (9.71) | 97.16 (1.48) | 96.68 (1.81) | -- | 21.78 (0.22) | 0.81 (0.20) | -- | -- | 0.21 | -- | 0.03 |
| | | **Avg.** | **55.26 (8.49)** | **93.01 (2.41)** | **91.84 (3.74)** | **84.10 (7.26)** | **20.14 (0.56)** | **1.03 (0.22)** | **0.24** | **0.13** | **0.27** | **0.30** | **0.07** |
| | 0.4 | <1 | 52.80 (14.28) | -- | -- | -- | 13.79 (1.84) | 0.75 (0.31) | 0.20 | 0.19 | -- | 0.47 | -- |
| | | 1 | 62.12 (4.19) | 80.36 (3.56) | 82.32 (4.33) | 68.37 (8.17) | 19.55 (0.58) | 2.02 (0.29) | 0.39 | 0.23 | 0.10 | 0.28 | 0.20 |
| | | 2 | 55.92 (7.99) | 77.54 (4.10) | 77.50 (5.79) | 61.16 (9.28) | 22.33 (0.55) | 2.83 (0.35) | 0.27 | 0.11 | 0.19 | 0.25 | 0.22 |
| | | 3 | 47.52 (8.22) | 76.66 (4.68) | 75.43 (6.14) | 56.39 (10.65) | 23.02 (0.66) | 2.63 (0.45) | 0.18 | 0.05 | 0.27 | 0.25 | 0.23 |
| | | 4 | 44.34 (7.39) | 75.94 (4.71) | 69.38 (7.29) | 53.58 (11.87) | 23.08 (0.54) | 2.30 (0.46) | 0.10 | 0.00 | 0.35 | 0.21 | 0.24 |
| | | 5 | 51.08 (10.30) | 75.82 (4.89) | 60.38 (6.58) | -- | 22.68 (0.52) | 1.81 (0.44) | -- | -- | 0.49 | -- | 0.24 |
| | | >5 | 78.16 (10.20) | 88.10 (3.26) | 86.04 (3.81) | -- | 22.56 (0.38) | 1.61 (0.36) | -- | -- | 0.22 | -- | 0.12 |
| | | **Avg.** | **55.99 (8.94)** | **79.07 (4.20)** | **75.17 (5.66)** | **59.88 (9.99)** | **21.00 (0.73)** | **1.99 (0.38)** | **0.23** | **0.12** | **0.27** | **0.29** | **0.21** |
| 30 | 0.3 | <1 | 53.20 (14.56) | -- | -- | -- | 14.15 (2.09) | 0.47 (0.20) | 0.21 | 0.22 | -- | 0.47 | -- |
| | | 1 | 60.90 (4.50) | 96.54 (1.73) | 97.17 (2.04) | 94.03 (5.03) | 20.63 (0.67) | 1.23 (0.21) | 0.43 | 0.30 | 0.10 | 0.29 | 0.03 |
| | | 2 | 56.92 (6.79) | 94.94 (2.14) | 95.11 (2.73) | 91.07 (5.25) | 24.06 (0.71) | 1.67 (0.25) | 0.32 | 0.18 | 0.18 | 0.26 | 0.05 |
| | | 3 | 50.26 (8.23) | 95.10 (2.19) | 95.29 (2.75) | 90.30 (6.12) | 26.49 (0.60) | 1.69 (0.26) | 0.25 | 0.14 | 0.25 | 0.25 | 0.05 |
| | | 4 | 48.04 (7.27) | 94.52 (2.30) | 94.26 (3.91) | 87.08 (7.46) | 28.29 (0.51) | 1.62 (0.24) | 0.17 | 0.00 | 0.31 | 0.21 | 0.05 |
| | | 5 | 62.56 (8.25) | 91.54 (2.93) | 86.81 (4.11) | -- | 27.97 (0.48) | 1.08 (0.18) | -- | -- | 0.37 | -- | 0.08 |
| | | >5 | 90.88 (5.82) | 98.50 (1.05) | 98.39 (1.11) | -- | 27.12 (0.53) | 0.83 (0.19) | -- | -- | 0.09 | -- | 0.02 |
| | | **Avg.** | **60.39 (7.92)** | **95.19 (2.06)** | **94.51 (2.77)** | **90.62 (5.96)** | **24.10 (0.80)** | **1.23 (0.22)** | **0.28** | **0.17** | **0.22** | **0.29** | **0.05** |
| | 0.4 | <1 | 53.68 (13.78) | -- | -- | -- | 14.23 (2.06) | 0.79 (0.34) | 0.21 | 0.20 | -- | 0.46 | -- |
| | | 1 | 64.34 (4.22) | 82.08 (3.56) | 82.87 (3.93) | 72.67 (8.12) | 21.15 (0.70) | 2.20 (0.24) | 0.41 | 0.27 | 0.10 | 0.25 | 0.18 |
| | | 2 | 58.06 (6.95) | 79.66 (4.25) | 78.36 (5.20) | 68.62 (10.27) | 25.31 (0.79) | 3.31 (0.42) | 0.30 | 0.15 | 0.18 | 0.24 | 0.20 |
| | | 3 | 51.86 (8.68) | 81.34 (4.70) | 78.93 (5.40) | 67.42 (10.19) | 27.90 (0.84) | 3.45 (0.53) | 0.24 | 0.11 | 0.26 | 0.22 | 0.19 |
| | | 4 | 48.66 (6.96) | 80.76 (4.03) | 76.91 (6.57) | 64.82 (11.79) | 29.82 (0.69) | 3.30 (0.48) | 0.16 | 0.00 | 0.31 | 0.20 | 0.19 |
| | | 5 | 63.06 (10.22) | 76.68 (4.57) | 64.48 (6.63) | -- | 29.20 (0.57) | 2.46 (0.50) | -- | -- | 0.37 | -- | 0.23 |
| | | >5 | 90.42 (6.18) | 92.50 (2.47) | 91.98 (2.72) | -- | 28.16 (0.60) | 1.90 (0.37) | -- | -- | 0.10 | -- | 0.08 |
| | | **Avg.** | **61.44 (8.14)** | **82.17 (3.93)** | **78.92 (5.07)** | **68.38 (10.09)** | **25.11 (0.89)** | **2.49 (0.41)** | **0.26** | **0.15** | **0.22** | **0.28** | **0.18** |

Table S6: Summary of analysis assessing activity specificity regarding identification accuracy, number of treated patients, overdose rate, underestimation rate, and overestimation rate of BF-BOLD design with 5-dose lattice. The true activity rates at MTD are 0.3 and 0.4, less than $\phi^a = 0.5$. The accuracy rate of OBD identification refers to the likelihood of no OBD being identified (i.e. specificity). All the other design characteristics, parameter settings, and analysis procedures are identical to those in Table S1.

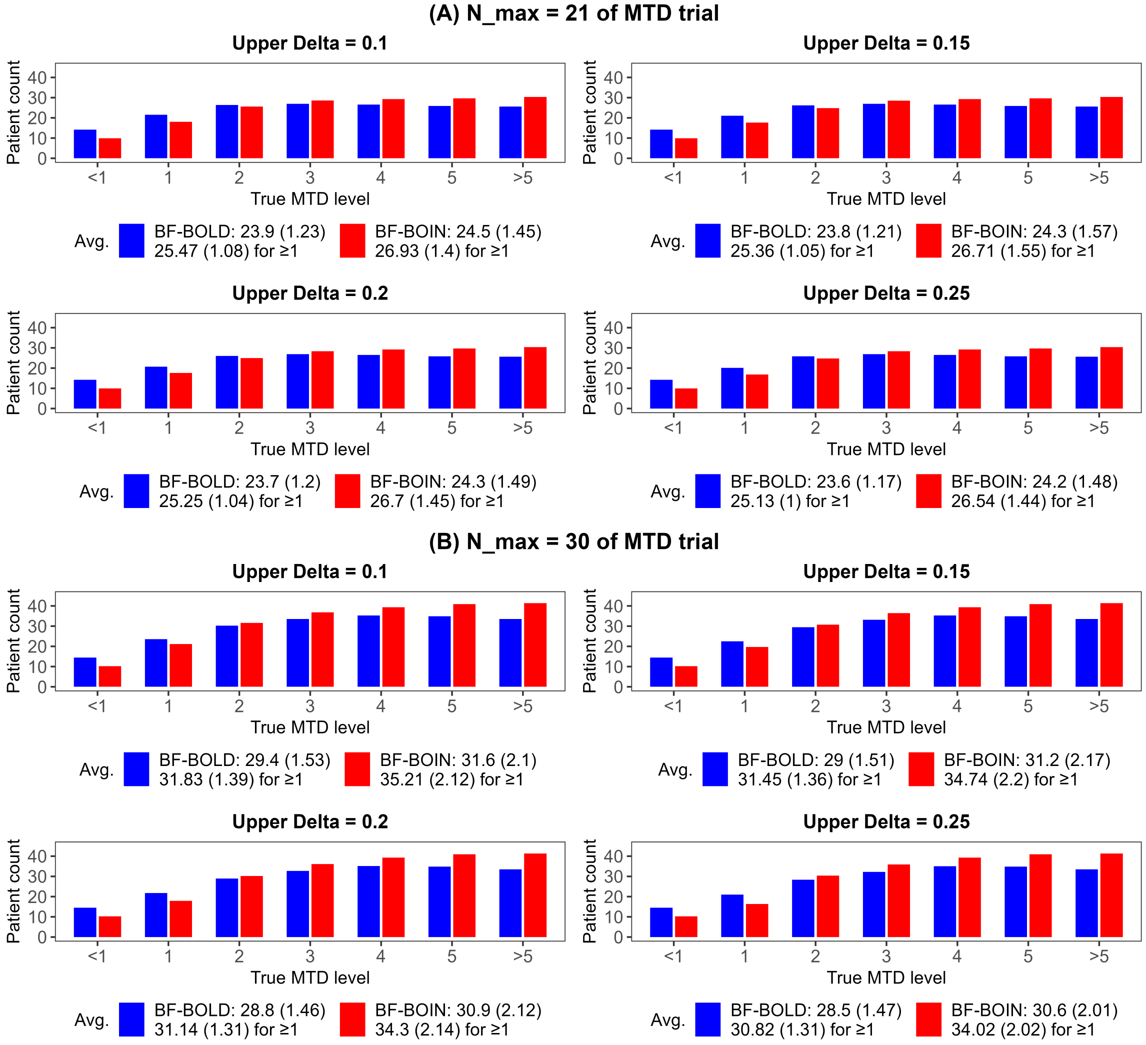


Figure S2: Bar charts illustrating comparative analysis of MTD efficiency (patient count with standard deviation) between BF-BOLD and BF-BOIN for 5-dose lattice, stratified by $N_{max}$ and upper $\delta$ of DLT rate, while OBD is assumed to be zero level below MTD (gap $= 0$) and a plateau of activity is present. All the other design characteristics and parameter settings are identical to those in Table S1. The legend denotes the average for all the 7 MTD levels (1st row) and for the MTD levels that are $\geq 1$ (2nd row).

| Gap (MTD minus OBD) | MTD level | Accuracy of identification (%) | | | | | Number of treated patients | | Overdose rate | |
|---|---|---|---|---|---|---|---|---|---|---|
| | | MTD | | OBD (BF-BOLD) | | | | | | |
| | | BF-BOLD | BF-BOIN | Overall | When identified MTD = true MTD | When identified MTD > true MTD | BF-BOLD | BF-BOIN | BF-BOLD | BF-BOIN |
| 0 | <1 | 58.40 (16.31) | 77.54 (12.98) | -- | -- | -- | 13.64 (2.27) | 9.65 (2.05) | 0.16 | 0.14 |
| | 1 | 74.48 (4.39) | 60.46 (1.96) | 80.22 (3.13) | 95.16 (2.56) | 64.43 (13.42) | 20.59 (0.62) | 17.09 (0.44) | 0.32 | 0.29 |
| | 2 | 66.76 (9.66) | 59.74 (12.80) | 46.36 (7.55) | 60.71 (6.49) | 43.16 (14.18) | 25.76 (0.76) | 24.85 (2.31) | 0.19 | 0.13 |
| | 3 | 69.72 (11.76) | 60.10 (15.22) | 45.62 (9.24) | 65.29 (5.58) | -- | 25.59 (0.82) | 27.61 (1.76) | -- | -- |
| | >3 | 95.38 (3.89) | 89.44 (8.43) | 57.76 (9.49) | 60.57 (9.75) | -- | 23.06 (1.54) | 27.00 (1.15) | -- | -- |
| | **Avg.** | **72.95 (9.20)** | **69.46 (10.28)** | **57.49 (7.35)** | **70.43 (6.09)** | **53.80 (13.80)** | **21.73 (1.20)** | **21.24 (1.54)** | **0.22** | **0.18** |
| | **(≥1)** | **76.59 (7.42)** | **67.44 (9.60)** | **57.49 (7.35)** | **70.43 (6.09)** | **53.80 (13.80)** | **23.75 (0.93)** | **24.14 (1.42)** | **0.26** | **0.21** |
| 1 | <1 | 58.40 (16.31) | 77.54 (12.98) | -- | -- | -- | 13.64 (2.27) | 9.65 (2.05) | 0.16 | 0.14 |
| | 1 | 74.48 (4.39) | 60.46 (1.96) | -- | -- | -- | 20.59 (0.62) | 17.09 (0.44) | 0.32 | 0.29 |
| | 2 | 66.64 (9.82) | 58.26 (13.52) | 69.44 (5.52) | 67.35 (5.79) | 48.49 (16.39) | 26.32 (0.75) | 25.28 (2.24) | 0.19 | 0.13 |
| | 3 | 70.00 (11.43) | 58.90 (13.52) | 45.98 (5.50) | 43.47 (6.17) | -- | 27.45 (0.68) | 28.75 (2.14) | -- | -- |
| | >3 | 94.82 (5.34) | 88.56 (9.11) | 61.54 (5.75) | 64.89 (4.57) | -- | 25.91 (0.59) | 28.25 (0.91) | -- | -- |
| | **Avg.** | **72.87 (9.46)** | **68.74 (10.22)** | **58.99 (5.59)** | **58.57 (5.51)** | **48.49 (16.39)** | **22.78 (0.98)** | **21.80 (1.56)** | **0.22** | **0.18** |
| | **(≥1)** | **76.49 (7.74)** | **66.55 (9.53)** | **58.99 (5.59)** | **58.57 (5.51)** | **48.49 (16.39)** | **25.07 (0.66)** | **24.84 (1.43)** | **0.25** | **0.21** |
| 2 | <1 | 58.40 (16.31) | 77.54 (12.98) | -- | -- | -- | 13.64 (2.27) | 9.65 (2.05) | 0.16 | 0.14 |
| | 1 | 74.48 (4.39) | 60.46 (1.96) | -- | -- | -- | 20.59 (0.62) | 17.09 (0.44) | 0.32 | 0.29 |
| | 2 | 66.64 (9.82) | 58.26 (13.52) | -- | -- | -- | 26.32 (0.75) | 25.28 (2.24) | 0.19 | 0.13 |
| | 3 | 69.48 (11.31) | 60.26 (13.21) | 53.00 (6.20) | 45.47 (7.09) | -- | 27.70 (0.72) | 29.02 (2.16) | -- | -- |
| | >3 | 95.54 (4.57) | 87.38 (8.77) | 44.36 (5.62) | 43.54 (5.62) | -- | 27.68 (0.28) | 29.18 (0.59) | -- | -- |
| | **Avg.** | **72.91 (9.28)** | **68.78 (10.09)** | **48.68 (5.91)** | **44.51 (6.36)** | -- | **23.19 (0.93)** | **22.04 (1.50)** | **0.22** | **0.18** |
| | **(≥1)** | **76.54 (7.52)** | **66.59 (9.37)** | **48.68 (5.91)** | **44.51 (6.36)** | -- | **25.57 (0.59)** | **25.14 (1.36)** | **0.25** | **0.21** |

Table S7: Summary of identification accuracy, number of treated patients and overdose rate of BF-BOLD and BF-BOIN design with 3-dose lattice for $N_{max} = 21$ and upper $\delta$ of true DLT rates $= 0.2$, where "Avg." denotes the average for all the 5 MTD levels and "($\geq$1)" denotes the average for the MTD levels that are $\geq 1$. All the other design characteristics and parameter settings are identical to those in Table S1.

| Gap (MTD minus OBD) | MTD level | Accuracy of identification (%) | | | | | Number of treated patients | | Overdose rate | |
|---|---|---|---|---|---|---|---|---|---|---|
| | | MTD | | OBD (BF-BOLD) | | | | | | |
| | | BF-BOLD | BF-BOIN | Overall | When identified MTD = true MTD | When identified MTD > true MTD | BF-BOLD | BF-BOIN | BF-BOLD | BF-BOIN |
| 0 | <1 | 58.38 (16.24) | 79.20 (11.98) | -- | -- | -- | 13.82 (2.33) | 9.83 (2.17) | 0.16 | 0.14 |
| | 1 | 75.88 (3.93) | 60.88 (2.89) | 81.46 (3.59) | 95.74 (2.74) | 66.37 (12.30) | 21.50 (0.77) | 18.73 (0.73) | 0.33 | 0.29 |
| | 2 | 67.96 (10.15) | 63.42 (14.06) | 47.74 (8.47) | 62.99 (5.45) | 38.89 (13.31) | 28.15 (0.97) | 29.90 (3.35) | 0.19 | 0.14 |
| | 3 | 71.66 (13.48) | 65.90 (15.56) | 47.06 (10.02) | 65.56 (5.77) | -- | 27.48 (0.98) | 31.84 (2.71) | -- | -- |
| | >3 | 96.34 (3.47) | 92.26 (7.01) | 60.12 (9.63) | 62.45 (10.02) | -- | 23.69 (1.72) | 29.14 (1.40) | -- | -- |
| | **Avg.** | **74.04 (9.45)** | **72.33 (10.30)** | **59.10 (7.93)** | **71.68 (5.99)** | **52.63 (12.80)** | **22.93 (1.36)** | **23.89 (2.07)** | **0.22** | **0.19** |
| | **(≥1)** | **77.96 (7.76)** | **70.62 (9.88)** | **59.10 (7.93)** | **71.68 (5.99)** | **52.63 (12.80)** | **25.21 (1.11)** | **27.40 (2.05)** | **0.26** | **0.22** |
| 1 | <1 | 58.38 (16.24) | 79.20 (11.98) | -- | -- | -- | 13.82 (2.33) | 9.83 (2.17) | 0.16 | 0.14 |
| | 1 | 75.88 (3.93) | 60.88 (2.89) | -- | -- | -- | 21.50 (0.77) | 18.73 (0.73) | 0.33 | 0.29 |
| | 2 | 68.54 (10.73) | 64.46 (14.47) | 70.22 (4.55) | 68.82 (4.79) | 47.07 (12.98) | 28.54 (0.91) | 29.64 (2.86) | 0.18 | 0.14 |
| | 3 | 70.68 (11.52) | 67.42 (14.91) | 46.82 (5.27) | 44.23 (5.67) | -- | 29.47 (0.83) | 33.42 (2.51) | -- | -- |
| | >3 | 96.22 (4.36) | 91.60 (7.27) | 62.06 (5.12) | 64.53 (4.93) | -- | 27.11 (0.73) | 30.44 (1.01) | -- | -- |
| | **Avg.** | **73.94 (9.36)** | **72.71 (10.30)** | **59.70 (4.98)** | **59.20 (5.13)** | **47.07 (12.98)** | **24.09 (1.11)** | **24.41 (1.86)** | **0.22** | **0.19** |
| | **(>1)** | **77.83 (7.63)** | **71.09 (9.89)** | **59.70 (4.98)** | **59.20 (5.13)** | **47.07 (12.98)** | **26.66 (0.81)** | **28.06 (1.78)** | **0.26** | **0.22** |
| 2 | <1 | 58.38 (16.24) | 79.20 (11.98) | -- | -- | -- | 13.82 (2.33) | 9.83 (2.17) | 0.16 | 0.14 |
| | 1 | 75.88 (3.93) | 60.88 (2.89) | -- | -- | -- | 21.50 (0.77) | 18.73 (0.73) | 0.33 | 0.29 |
| | 2 | 68.54 (10.73) | 64.46 (14.47) | -- | -- | -- | 28.54 (0.91) | 29.64 (2.86) | 0.18 | 0.14 |
| | 3 | 71.34 (11.97) | 67.02 (14.88) | 53.62 (6.45) | 46.00 (6.78) | -- | 29.77 (0.74) | 33.42 (2.52) | -- | -- |
| | >3 | 95.70 (4.11) | 92.30 (6.18) | 43.82 (5.64) | 43.22 (5.58) | -- | 28.85 (0.66) | 31.73 (0.87) | -- | -- |
| | **Avg.** | **73.97 (9.39)** | **72.77 (10.08)** | **48.72 (6.04)** | **44.61 (6.18)** | -- | **24.50 (1.08)** | **24.67 (1.83)** | **0.22** | **0.19** |
| | **(>1)** | **77.87 (7.68)** | **71.17 (9.60)** | **48.72 (6.04)** | **44.61 (6.18)** | -- | **27.16 (0.77)** | **28.38 (1.74)** | **0.26** | **0.22** |

Table S8: Summary of identification accuracy, number of treated patients and overdose rate of BF-BOLD and BF-BOIN design with 3-dose lattice for $N_{max} = 30$ and upper $\delta$ of true DLT rates $= 0.2$, where "Avg." denotes the average for all the 5 MTD levels and "($\geq 1$)" denotes the average for the MTD levels that are $\geq 1$. All the other design characteristics and parameter settings are identical to those in Table S1.

| Gap (MTD minus OBD) | MTD level | Overdose rate | | Underestimation rate | | | | Overestimation rate | | | |
|---|---|---|---|---|---|---|---|---|---|---|---|
| | | | | MTD | | OBD | | MTD | | OBD | |
| | | BF-BOLD | BOLD-1b | BF-BOLD | BOLD-1b | BF-BOLD | BOLD-1b | BF-BOLD | BOLD-1b | BF-BOLD | BOLD-1b |
| 0 | <1 | 0.19 | 0.20 | -- | -- | -- | -- | 0.45 | 0.34 | -- | -- |
| | 1 | 0.37 | 0.40 | 0.11 | 0.13 | 0.15 | 0.14 | 0.21 | 0.29 | 0.07 | 0.12 |
| | 2 | 0.25 | 0.30 | 0.19 | 0.19 | 0.50 | 0.49 | 0.21 | 0.26 | 0.06 | 0.09 |
| | 3 | 0.18 | 0.22 | 0.28 | 0.27 | 0.55 | 0.54 | 0.22 | 0.24 | 0.07 | 0.09 |
| | 4 | 0.11 | 0.14 | 0.34 | 0.37 | 0.61 | 0.63 | 0.20 | 0.18 | 0.06 | 0.06 |
| | 5 | -- | -- | 0.43 | 0.49 | 0.67 | 0.72 | -- | -- | -- | -- |
| | >5 | -- | -- | 0.15 | 0.24 | 0.63 | 0.73 | -- | -- | -- | -- |
| | **Avg.** | **0.22** | **0.25** | **0.25** | **0.28** | **0.52** | **0.54** | **0.26** | **0.26** | **0.07** | **0.09** |
| | **(≥1)** | **0.23** | **0.26** | **0.25** | **0.28** | **0.52** | **0.54** | **0.21** | **0.24** | **0.07** | **0.09** |
| 1 | <1 | 0.19 | 0.20 | -- | -- | -- | -- | 0.45 | 0.34 | -- | -- |
| | 1 | 0.37 | 0.40 | 0.11 | 0.13 | -- | -- | 0.21 | 0.29 | -- | -- |
| | 2 | 0.24 | 0.30 | 0.20 | 0.19 | 0.03 | 0.03 | 0.21 | 0.26 | 0.29 | 0.34 |
| | 3 | 0.17 | 0.22 | 0.29 | 0.27 | 0.35 | 0.35 | 0.21 | 0.24 | 0.22 | 0.28 |
| | 4 | 0.11 | 0.14 | 0.35 | 0.37 | 0.40 | 0.42 | 0.19 | 0.18 | 0.22 | 0.23 |
| | 5 | -- | -- | 0.44 | 0.51 | 0.46 | 0.49 | -- | -- | 0.17 | 0.16 |
| | >5 | -- | -- | 0.15 | 0.22 | 0.50 | 0.58 | -- | -- | -- | -- |
| | **Avg.** | **0.22** | **0.25** | **0.26** | **0.28** | **0.35** | **0.37** | **0.26** | **0.26** | **0.23** | **0.25** |
| | **(≥1)** | **0.22** | **0.26** | **0.26** | **0.28** | **0.35** | **0.37** | **0.21** | **0.24** | **0.23** | **0.25** |
| 2 | <1 | 0.19 | 0.20 | -- | -- | -- | -- | 0.45 | 0.34 | -- | -- |
| | 1 | 0.37 | 0.40 | 0.11 | 0.13 | -- | -- | 0.21 | 0.29 | -- | -- |
| | 2 | 0.24 | 0.30 | 0.20 | 0.19 | -- | -- | 0.21 | 0.26 | -- | -- |
| | 3 | 0.17 | 0.22 | 0.28 | 0.27 | 0.01 | 0.01 | 0.23 | 0.24 | 0.41 | 0.47 |
| | 4 | 0.10 | 0.14 | 0.36 | 0.36 | 0.32 | 0.31 | 0.19 | 0.18 | 0.33 | 0.38 |
| | 5 | -- | -- | 0.45 | 0.49 | 0.33 | 0.37 | -- | -- | 0.33 | 0.33 |
| | >5 | -- | -- | 0.16 | 0.24 | 0.36 | 0.42 | -- | -- | 0.27 | 0.26 |
| | **Avg.** | **0.22** | **0.25** | **0.26** | **0.28** | **0.25** | **0.28** | **0.26** | **0.26** | **0.33** | **0.36** |
| | **(≥1)** | **0.22** | **0.26** | **0.26** | **0.28** | **0.25** | **0.28** | **0.21** | **0.24** | **0.33** | **0.36** |

Table S9: Comparative analysis of BF-BOLD vs. BOLD-1b with a 5-dose model, for overdose rate, underestimation rate (MTD and OBD), and over estimation rate (MTD and OBD). A random number are backfilled (1 to 3) in BF-BOLD, while for BOLD-1b, 10 patients are treated at the identified MTD after using BOLD, with no backfilling. $N_{max}$ values are set to be 24 for BF-BOLD and 21 for BOLD-1b, respectively, ensuring that their overall numbers of treated patients are closely aligned. "Avg." denotes the average for all the 5 MTD levels and "($\geq$1)" denotes the average for the MTD levels that are $\geq 1$. All the other design characteristics and parameter settings are identical to those in Table S1.
